# The Estimated Time-Varying Reproduction Numbers during the Ongoing Pandemic of the Coronavirus Disease 2019 (COVID-19) in 12 Selected Countries outside China

**DOI:** 10.1101/2020.05.10.20097154

**Authors:** Fu-Chang Hu

## Abstract

**Background:** How can we anticipate the progression of the ongoing pandemic of the coronavirus disease 2019 (COVID-19)? As a measure of transmissibility, we aimed to estimate concurrently the time-varying reproduction number, *R*_0_(*t*), over time during the COVID-19 pandemic for each of the following 12 heavily-attacked countries: Singapore, South Korea, Japan, Iran, Italy, Spain, Germany, France, Belgium, United Kingdom, the United States of America, and South Africa.

**Methods:** We downloaded the publicly available COVID-19 pandemic data from the `WHO COVID-19 Dashboard` website (https://covid19.who.int/) for the duration of January 11, 2020 and May 1, 2020. Then, we specified two plausible distributions of serial interval to apply the novel estimation method implemented in the incidence and `EpiEstim` packages to the data of daily new confirmed cases for robustly estimating *R*_0_(*t*) in the R software.

**Results:** We plotted the epidemic curves of daily new confirmed cases for the 12 selected countries. A clear peak of the epidemic curve appeared in 10 of the 12 selected countries at various time points, and then the epidemic curve declined gradually. However, the United States of America and South Africa happened to have two or more peaks and their epidemic curves either reached a plateau or still climbed up. Almost all curves of the estimated *R*_0_(*t*) monotonically went down to be less than or close to 1.0 up to April 30, 2020 except Singapore, South Korea, Japan, Iran, and South Africa, of which the curves surprisingly went up and down at various time periods during the COVID-19 pandemic. Finally, the United States of America and South Africa were the two countries with the approximate *R*_0_(*t*) ≥ 1.0 at the end of April, and thus they were now facing the harshest battles against the coronavirus among the 12 selected countries. By contrast, Spain, Germany, and France with smaller values of the estimated *R*_0_(*t*) were relatively better than the other 9 countries.

**Conclusion:** Seeing the estimated *R*_0_(*t*) going downhill speedily is more informative than looking for the drops in the daily number of new confirmed cases during an ongoing epidemic of infectious disease. We urge public health authorities and scientists to estimate *R*_0_(*t*) routinely during an epidemic of infectious disease and to report *R*_0_(*t*) daily to the public until the end of the epidemic.

## Introduction

The coronavirus disease 2019 (COVID-19) is now known to be caused by the severe acute respiratory syndrome coronavirus 2 (SARS-CoV-2). The COVID-19 was first reported to the World Health Organization (WHO) from Wuhan City, Hubei Province, China on December 31, 2019. China locked down Wuhan on January 23, 2020 to prevent the coronavirus from spreading out to the other provinces of China and the other countries outside China. The WHO officially declared the COVID-19 epidemic as a Public Health Emergency of International Concern on January 30, 2020. After drastic control measures had been implemented with a high cost to combat the COVID-19 epidemic, President Xi Jinping of China made his first visit to Wuhan on March 10, 2020 to show that China turned the corner on coronavirus. China announced no new locally transmitted cases on March 19, 2020, and then ended its lockdown of Wuhan on April 8, 2020. Nevertheless, the WHO described coronavirus as a pandemic on March 11, 2020 due to its rapid and wide spread over the world outside China. Traveling oversea has dramatically shortened the social distances between countries worldwide, and thus the coronavirus spreads out more quickly and widely than ever before. At the same time, public panic spreads out even more drastically through social and news media. The epidemic of such a novel infectious disease has caused big damages to human health, medical facilities, economy, and society in many countries. Once the epidemic is likely to occur, appropriate control tactics and measures according to the source-transmission-host theory, including the nonpharmaceutical interventions (NPI) and pharmaceutical interventions (PI), should be implemented timely to lower such damages (see Appendix).^1^ In China, prompt and aggressive actions had been taken since the very early phase of the epidemic.^2^ In the other counties, areas, or territories such as Taiwan, Singapore, Thailand, Philippines, Japan, South Korea, Iran, Italy, Spain, France, Germany, United Kingdom, the United States of America, Australia, and South Africa, the sooner the proper actions were taken against the local spread of the coronavirus, the smaller the size of the COVID-19 epidemic and its damage would likely be. Since the coronavirus is highly transmittable by droplets, more stringent control measures such as fast and mass testing, rigorous contact tracing, large-scale isolations, mandatory quarantines, traveling restrictions or bans, border closing, social distancing, school closings, stay-at-home orders, curfews, and long-term lockdowns may be needed to contain the epidemic ultimately. Almost the whole world is now in the battle against the coronavirus as the global numbers of confirmed COVID-19 cases and deaths are soaring speedily.^3,4^

In such a disaster, various sectors of society should collaborate immediately to tackle the problem under the guidance of the government. Scientists and technology experts may try all the available tools to help the government fight the epidemic. Yet, how can we anticipate the progression of the ongoing COVID-19 pandemic in a city, province, or country? The *basic reproduction number, R*_0_, for an infectious disease is the expected number of new cases infected directly from the index case in a susceptible population. As a measure of transmissibility, it can tell us how quickly an infectious disease spreads out in various stages of the epidemic.^1^ Most importantly, *R*_0_ can be used to assess the effectiveness of the implemented control measures and to explore the possibility of pathogen mutations in an epidemic. When *R*_0_ < 1.0 persistently, the epidemic would be damped down soon.^1^ In essence, *R*_0_ is the driving force behind the epidemic curve. Its value usually varies over time during an epidemic. Hence, as biostatisticians, we had analyzed the epidemic data of China to estimate the time-varying reproduction number, *R*_0_(*t*), and reported the analysis result.^2^ Then, we started this investigation on April 4, 2020 with the aim to estimate concurrently the time-varying reproduction number, *R*_0_(*t*), over time during the ongoing COVID-19 pandemic for each of the following 12 heavily-attacked countries in different continents: (1) Singapore, (2) Republic of Korea (i.e., South Korea), (3) Japan, (4) Islamic Republic of Iran (i.e., Iran), (5) Italy, (6) Spain, (7) Germany, (8) France, (9) Belgium, (10) United Kingdom, (11) the United States of America, and (12) South Africa.

## Methods

### Epidemic data

To collect all individual patient data, including personal contact history, during an epidemic is a tough and crucial task. As revealed in the report of the Novel Coronavirus Pneumonia Emergency Response Epidemiology Team, great efforts have previously been made in China to build up China’s Infectious Disease Information System, which is very helpful in the management of nationwide patient data during this epidemic of a large size.^5^ Such highly confidential and miscellaneous data are not made available in any country to unauthorized researchers during the COVID-19 pandemic. Nevertheless, since January 11, 2020, the numbers of new confirmed cases, cumulative confirmed cases, new deaths, and cumulative deaths of the counties, areas, and territories in the world have been announced daily by the WHO on its official websites.^3,4^ Besides, the newly created and daily updated COVID-19 pandemic data R package, data2019nCoV, collects the COVID-19 pandemic data by transcribing or compiling the available data from official sources, including the WHO Situation Reports,^4^ on its website (https://github.com/eebrown/data2019nCoV). Thus, we downloaded the publicly available COVID-19 pandemic data (“WHO-COVID-19-global-data.csv”) from the WHO COVID-19 Dashboard website (https://covid19.who.int/),^3^ for the duration of January 11, 2020 and May 1, 2020 (Last Updated: 2020/5/1, 3:00 PM CEST) in this study.

### Epidemic analysis

Statistical analysis was performed using the R 3.6.3 software (R Foundation for Statistical Computing, Vienna, Austria). The distributional properties of daily counts of new confirmed cases were presented by the number of days (n), total number, mean, standard deviation (SD), minimum, the first quartile (Q1), median, the third quartile (Q3), and maximum for each of the 12 selected countries outside China. Instead of developing any advanced methods specific for this pandemic, we tried to find an available easy-to-use tool to monitor the progress of the ongoing COVID-19 pandemic as soon as possible. As listed on the Comprehensive R Archive Network (CRAN) (https://cran.r-project.org/), several R packages might be used to compute basic reproduction numbers of an epidemic in R, including argo, epibasix, EpiCurve, EpiEstim, EpiILM, EpiILMCT, epimdr,^6^ epinet, epiR, EpiReport, epitools, epitrix, incidence, mem, memapp, R0, and surveillance. We chose the incidence (version 1.7.0) and EpiEstim (version 2.2-1) packages to estimate *R*_0_(*t*) in R during the ongoing COVID-19 pandemic for the 12 selected countries outside China due to their methodological soundness and computational simplicity for rapid analysis.^7,8^ The R code was listed in the Supplementary Appendix of our previous study of China^2^ for check and re-uses.

First, we laid out the conceptual framework below. In an epidemic of infectious disease, any susceptible subject who becomes a patient usually goes through the following three dynamic stages: infection, development of symptoms, and diagnosis of the disease. Theoretically, to estimate *R*_0_ or *R*_0_(*t*), we need the information about the distribution of *generation time* (GT), which is the time interval between the infection of the index case and the infection of the next case infected directly from the index case.^8^ Yet, the time of infection is most likely unavailable or inaccurate, and thus investigators collect the data about the distribution of *serial interval* (SI) instead, which is the time interval between the symptom onset of the index case and the symptom onset of the next case infected directly from the index case.^8^ Nevertheless, the data of symptom onset are not publically available and almost always have the problem of delayed reporting in any ongoing epidemic of infectious disease because they are usually recorded at diagnosis.^5^ Hence, we took a common approach in statistics to tackle this problem by specifying the best plausible distributions of SI according to the results obtained from previous studies of similar epidemics, and then applied the novel estimation method implemented in the EpiEstim package to the data of daily new confirmed cases in practice.^7,8^

Next, we considered two plausible scenarios for studying the ongoing COVID-19 pandemic in the 12 selected countries outside China. The estimate_R function of the EpiEstim package assumes a Gamma distribution for SI by default to approximate the infectivity profile.^7^ Technically, the transmission of an infectious disease is modeled with a Poisson process in the EpiEstim package.^7,8^ When we choose a Gamma prior distribution for SI, the Bayesian statistical inference leads to a simple analytical expression for the Gamma posterior distribution of *R*_0_(*t*).^8^ In the first scenario, we specified the mean (SD) of the Gamma distribution for SI to be 8.4 (3.8) days to mimic the 2003 epidemic of the severe acute respiratory syndrome (SARS) in Hong Kong.^8^ Then, in the second scenario, we specified the mean (SD) of the Gamma distribution for SI to be 2.6 (1.5) days to mimic the 1918 pandemic of influenza in Baltimore, Maryland.^8^ According to the current understanding, the transmissibility of COVID-19 was higher than SARS but lower than influenza.^1^ Hence, even though we did not know the true distribution(s) of SI for the ongoing pandemic of COVID-19 in the 12 selected countries outside China, these two plausible scenarios helped us catch the behavior pattern of this pandemic along with the time evolution.

## Results

### Epidemic data

We subtracted one day from the reporting dates in the COVID-19 pandemic data of WHO to obtain the “dates” on which the daily new confirmed cases actually occurred. Then, we computed the sample statistics of the daily new confirmed cases for the 12 selected countries over the time period from January 10, 2020 to April 30, 2020 in Table 1. The starting date of the COVID-19 epidemic differed among the 12 countries so that the sample size (i.e., number of days) varied from 58 (South Africa) to 109 (Japan). All the daily new confirmed cases in the 12 selected countries were laboratory confirmed, but the number of daily new confirmed cases included both domestic and repatriated cases according to the description of WHO.^4^

**Table 1.** Descriptive statistics for the epidemic data of the coronavirus disease 2019 (COVID-19) in the 12 selected countries outside China from January 11, 2020 to May 1, 2020.

| Country | Number of Days | Total Number of Confirmed Cases | Daily New Confirmed Cases |  |  |  |  |  |  |
| --- | --- | --- | --- | --- | --- | --- | --- | --- | --- |
|  |  |  | Mean | SD | Minimum | Q1 | Median | Q3 | Maximum |
| **1. Singapore** | 100 | **16169** | **161.69** | 308.53 | 0 | 3.00 | 12.00 | 82.75 | **1426** |
| **2. South Korea** | 104 | **10774** | **103.60** | 166.73 | 0 | 1.00 | 27.00 | 104.25 | **813** |
| **3. Japan** | 109 | **14281** | **131.02** | 189.32 | 0 | 1.00 | 29.00 | 206.00 | **743** |
| **4. Iran** | 73 | **94640** | **1296.44** | 874.34 | 0 | 881.00 | 1192.00 | 1657.00 | **3186** |
| **5. Italy** | 94 | **203591** | **2165.86** | 2052.45 | 0 | 1.00 | 2173.50 | 3823.50 | **6557** |
| **6. Spain** | 92 | **212917** | **2314.32** | 2780.08 | 0 | 0 | 1045.50 | 4193.50 | **9222** |
| **7. Germany** | 95 | **159119** | **1674.94** | 2057.43 | 0 | 0.50 | 693.00 | 2843.50 | **7324** |
| **8. France** | 99 | **127066** | **1283.50** | 1664.70 | 0 | 0 | 385.00 | 2058.00 | **7500** |
| **9. Belgium** | 88 | **47859** | **543.85** | 628.52 | 0 | 0 | 242.00 | 1052.50 | **2454** |
| **10. United Kingdom** | 92 | **165225** | **1795.92** | 2239.15 | 0 | 0.75 | 229.50 | 4258.25 | **8719** |
| **11. United States of America** | 103 | **1035353** | **10051.97** | 13361.29 | 0 | 0 | 19.00 | 24274.50 | **38509** |
| **12. South Africa** | 58 | **5647** | **97.36** | 98.76 | 0 | 18.00 | 69.50 | 144.50 | **373** |

**Abbreviation:** Q1 = the first quartile, Q3 = the third quartile, and SD = standard deviation.

As expected, the larger the mean value of daily new confirmed cases, the bigger the size of the corresponding SD. Spain, France, Belgium, and the United States of America had zero-valued Q1’s, indicating that they had zero daily new confirmed case in the early phase of the pandemic for more than 25 days but the number of daily new confirmed cases were soaring quickly after then. These sample statistics revealed the magnitudes of the ongoing COVID-19 pandemic in each of the 12 selected countries over the study period. Surprisingly, the United States of America had a total of 1,035,353 new confirmed cases accumulated over 103 days up to April 30, 2020 and it became the severest epidemic area in the world.

### Epidemic analysis

First, we plotted the epidemic curves of daily new confirmed cases for each of the 12 selected countries in Figures 1-1, 2-1, …, 12-1 respectively. A clear peak of the epidemic curve appeared in 10 of the 12 selected countries at various time points, and then the epidemic curve declined gradually. However, the United States of America and South Africa happened to have two or more peaks and their epidemic curves either reached a plateau or still climbed up.

**Figure 1-1.**
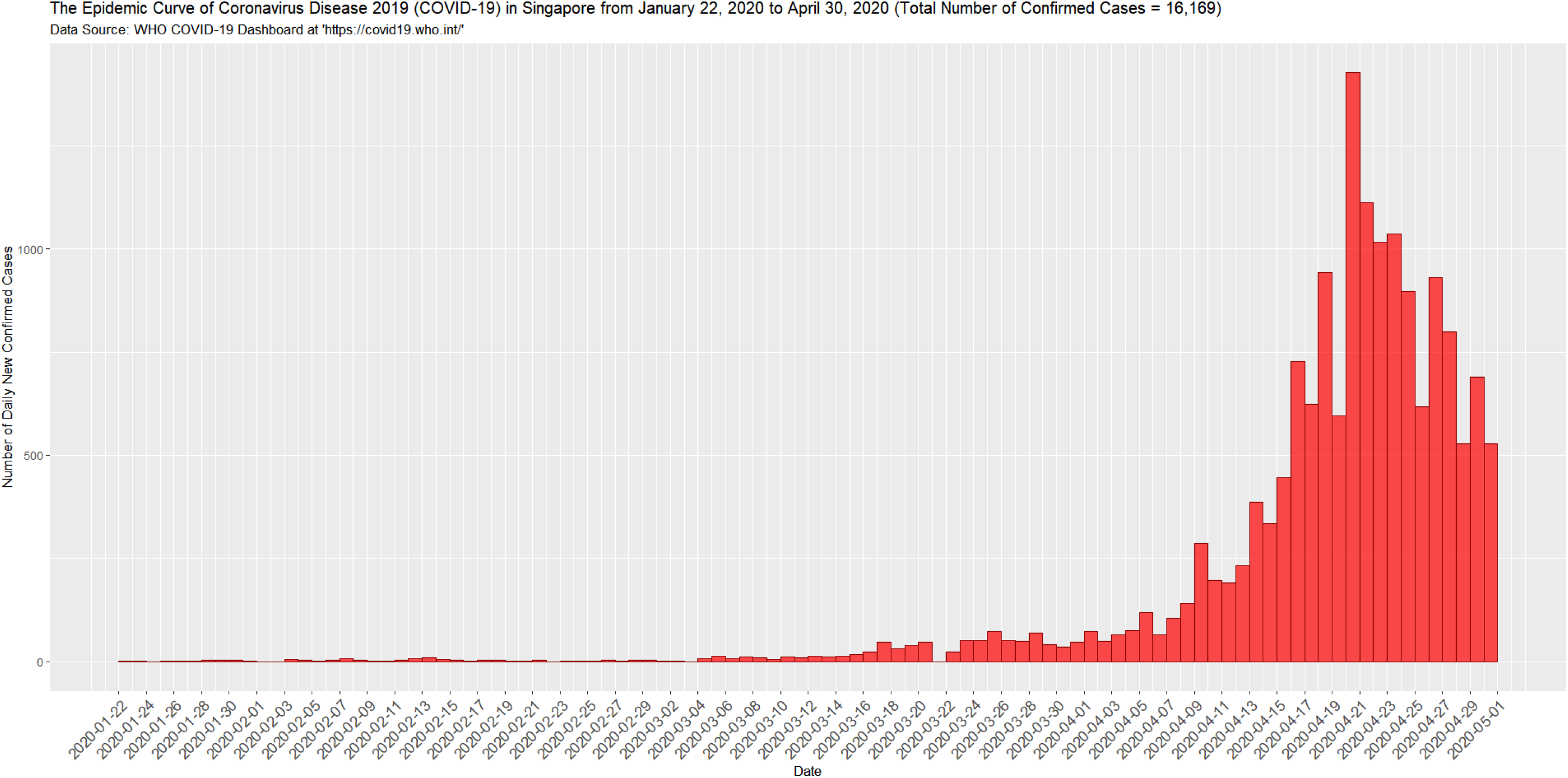
The epidemic curve of the coronavirus disease 2019 (COVID-19) in Singapore from January 22, 2020 to April 30, 2020.

**Figure 1-2.**
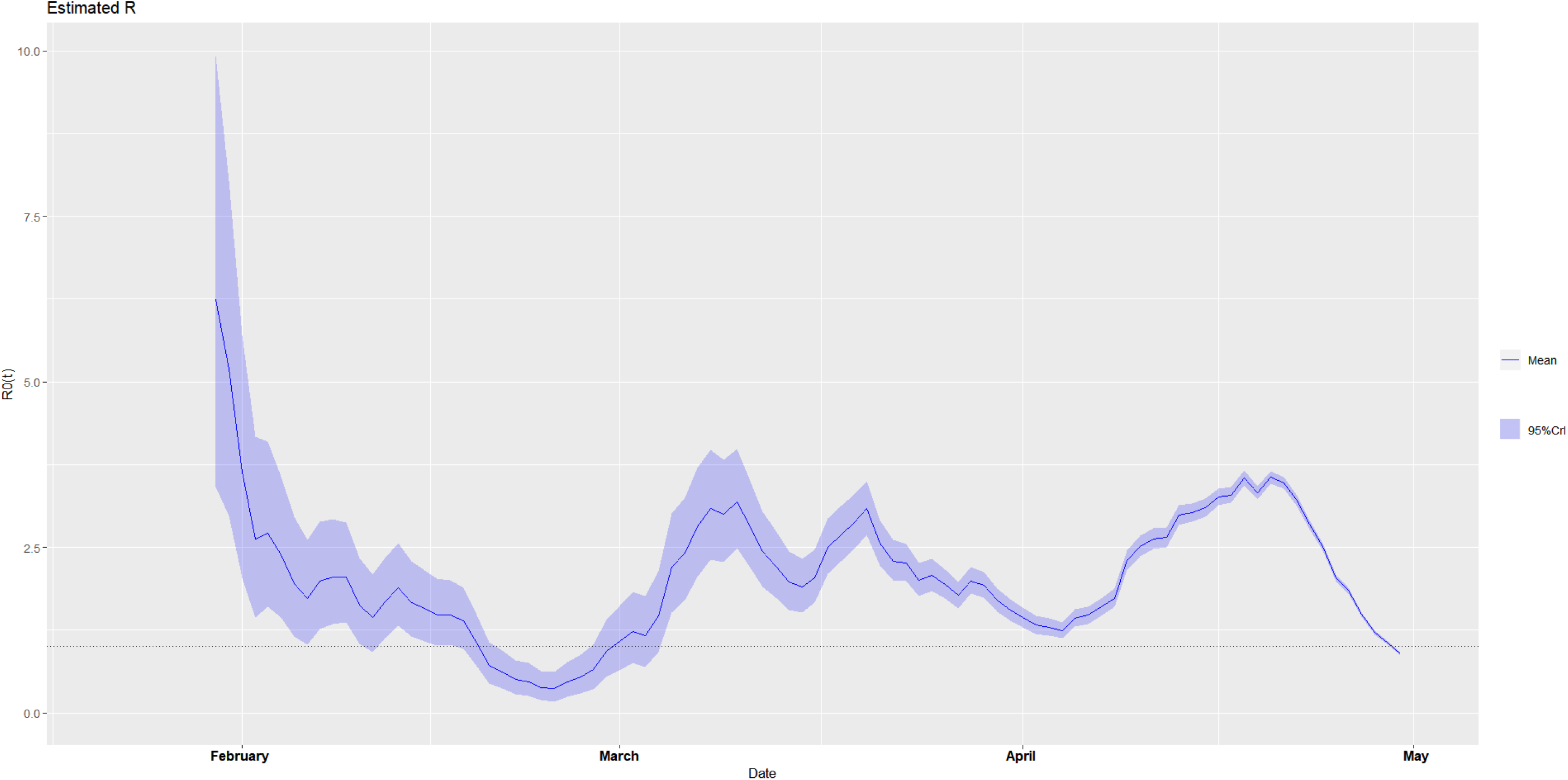
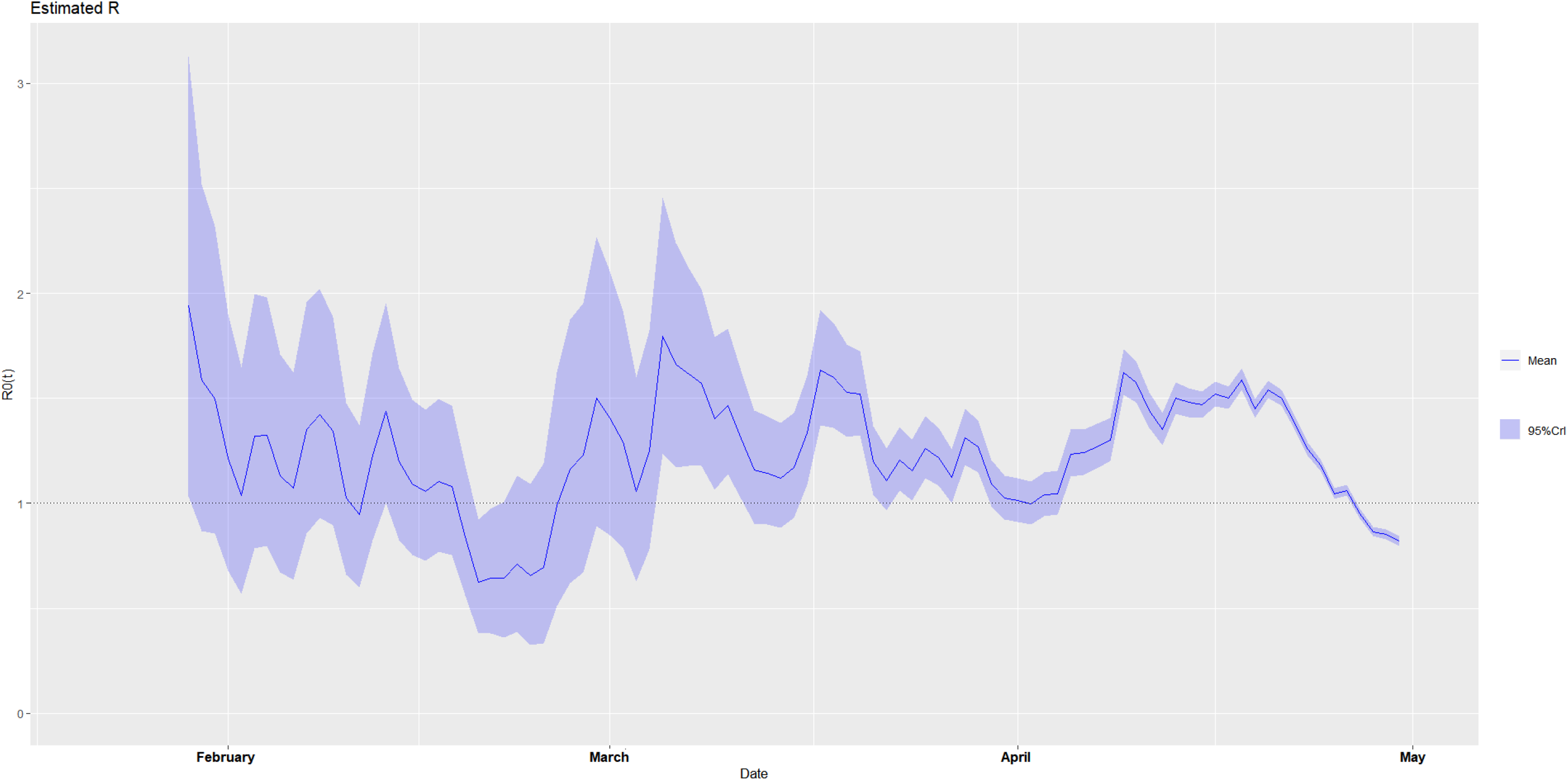
The estimated time-varying reproduction number during the ongoing pandemic of the coronavirus disease 2019 (COVID-19) in Singapore from January 22, 2020 to April 30, 2020 under two scenarios. **A. Scenario 1:** We specified the mean (SD) of the Gamma distribution of serial interval (SI) to be 8.4 (3.8) days to mimic the 2003 epidemic of the severe acute respiratory syndrome (SARS) in Hong Kong.^7,8^ **B. Scenario 2:** We specified the mean (SD) of the Gamma distribution of serial interval (SI) to be 2.6 (1.5) days to mimic the 1918 pandemic of influenza in Baltimore, Maryland.^7,8^

**Figure 2-1.**
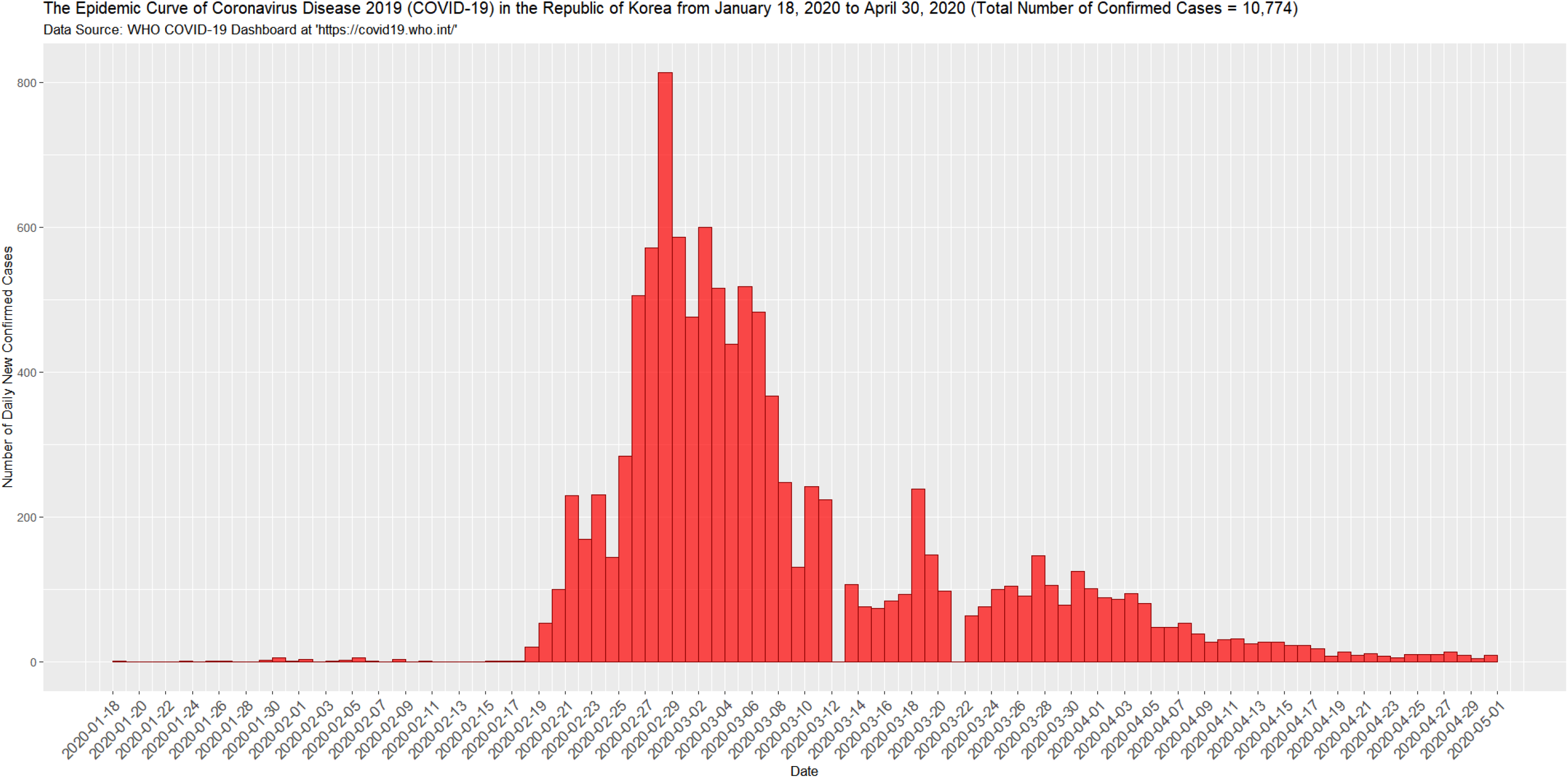
The epidemic curve of the coronavirus disease 2019 (COVID-19) in South Korea from January 18, 2020 to April 30, 2020.

Next, under the specified two plausible scenarios (**A** and **B**), we plotted the daily estimates of the time-varying reproduction numbers, *R*_0_(*t*), over sliding weekly windows,^7,8^ for each of the 12 selected countries in Figures 1-2A and 1-2B, 2-2A and 2-2B,…, and 12-2A and 12-2B respectively. On any given day of the epidemic curve, *R*_0_(*t*) was estimated for the weekly window ending on that day.^7,8^ The estimated *R*_0_(*t*) was not shown from the very beginning of the epidemic curve because precise estimation was not possible in that period.^8^ Specifically, the blue lines showed the posterior means of *R*_0_(*t*), the grey zones represented the 95% credible intervals (CrI), and the horizontal black dashed lines indicated the threshold value of *R*_0_ = 1.0.^7,8^ Since the repatriated cases were not separated from the domestic cases in the COVID pandemic data of WHO,^3,4^ they caused the problem of over-estimating *R*_0_(*t*) especially in the early phase of the COVID-19 pandemic, but they would have less and less effect on the estimation of *R*_0_(*t*) as time elapsed with traveling restrictions and bans worldwide. Almost all curves of the estimated *R*_0_(*t*) monotonically went down to be less than or close to 1.0 up to April 30, 2020, except Singapore, South Korea, Japan, Iran, and South Africa, of which the curves surprisingly went up and down at various time periods during the COVID-19 pandemic. Nevertheless, Singapore took stringent control measures lately, including a strengthened restriction of the population mobility, to contain the second and third waves of the COVID-19 epidemic. In particular, the effectiveness of these stringent control measures had led to a sharp drop in the estimated *R*_0_(*t*) to bring it down to below 1.0 in the last two weeks of April. This was a rarely seen victory in the places outside China, and thus it would be very encouraging to the countries with gently sloped curves for the last several weeks before April 30, 2020 such as South Korea, Japan, Iran, Italy, Spain, Germany, France, Belgium, United Kingdom, and the United States of America.

**Figure 2-2.**
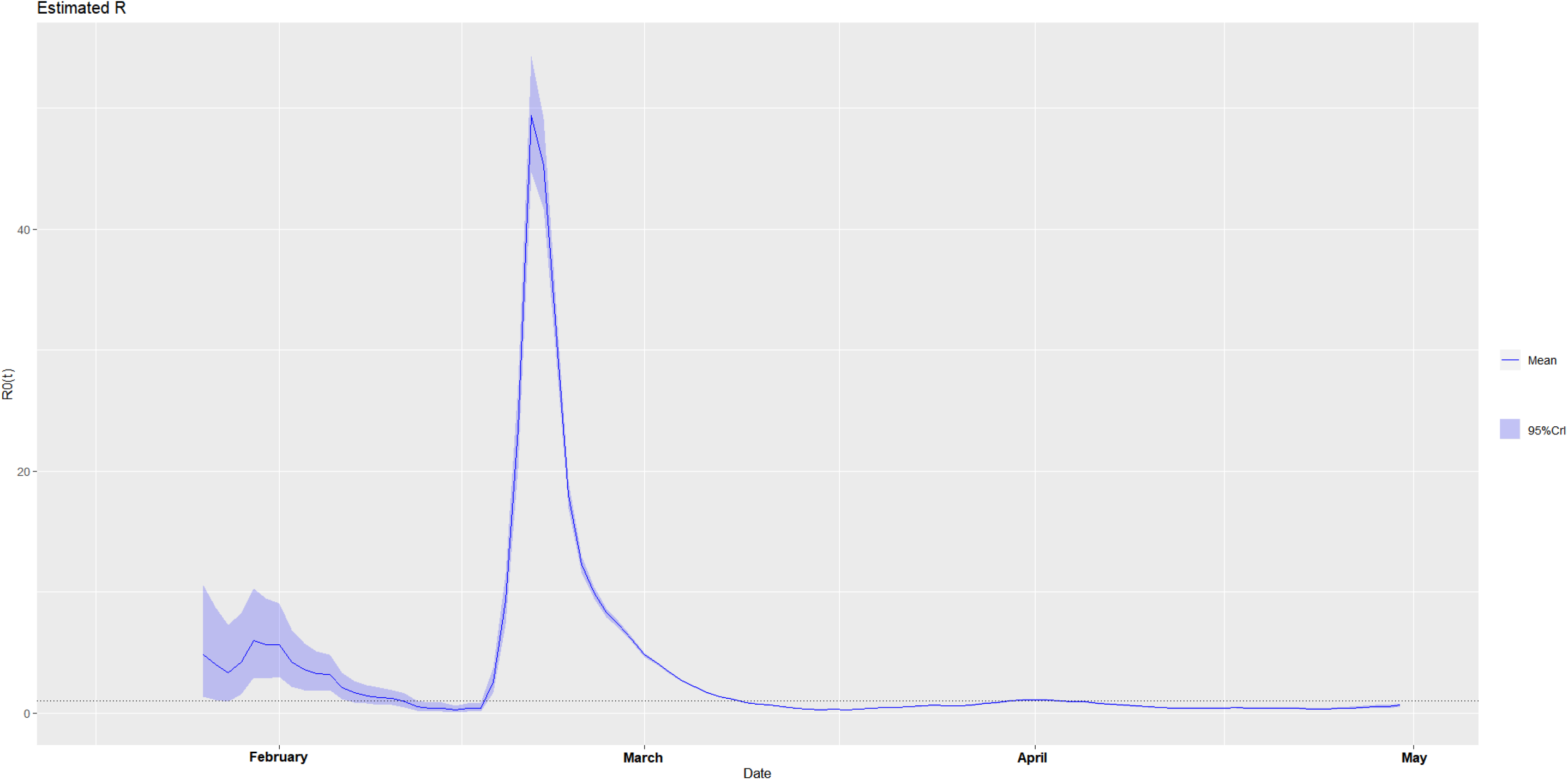
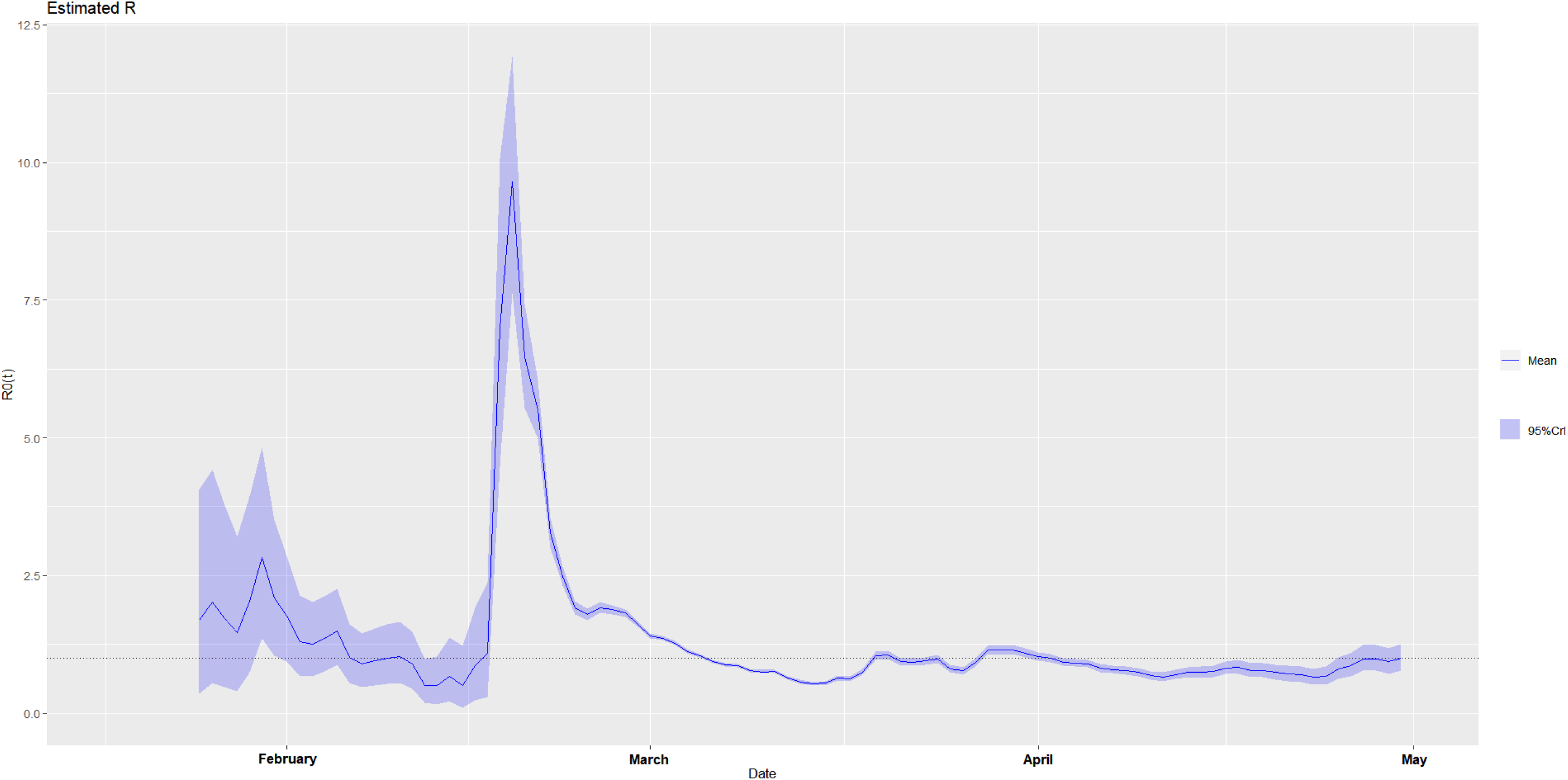
The estimated time-varying reproduction number during the ongoing pandemic of the coronavirus disease 2019 (COVID-19) in South Korea from January 18, 2020 to April 30, 2020 under two scenarios. **A. Scenario 1:** We specified the mean (SD) of the Gamma distribution of serial interval (SI) to be 8.4 (3.8) days to mimic the 2003 epidemic of the severe acute respiratory syndrome (SARS) in Hong Kong.^7,8^ **B. Scenario 2:** We specified the mean (SD) of the Gamma distribution of serial interval (SI) to be 2.6 (1.5) days to mimic the 1918 pandemic of influenza in Baltimore, Maryland.^7,8^

**Figure 3-1.**
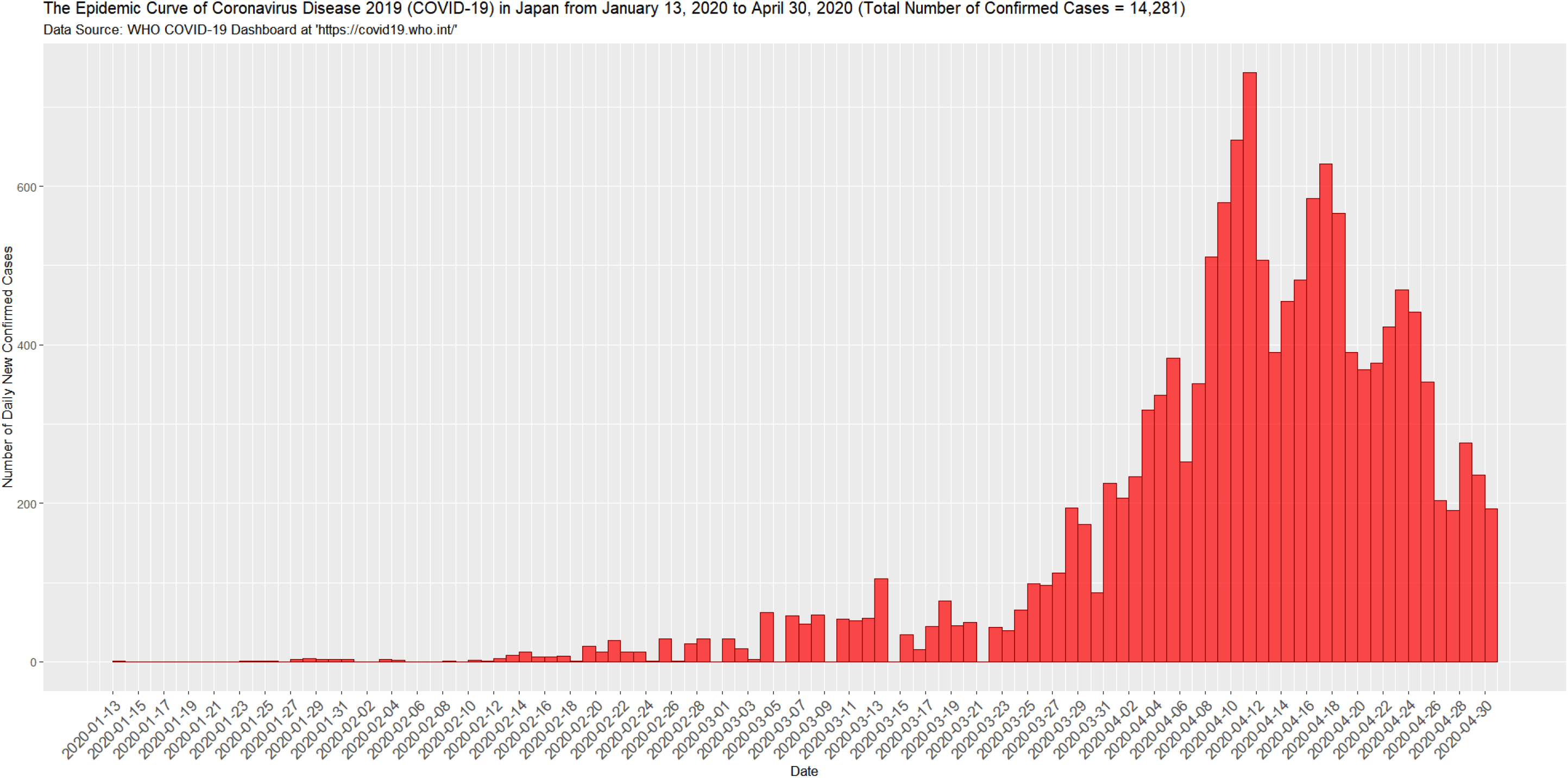
The epidemic curve of the coronavirus disease 2019 (COVID-19) in Japan from January 13, 2020 to April 30, 2020.

**Figure 3-2.**
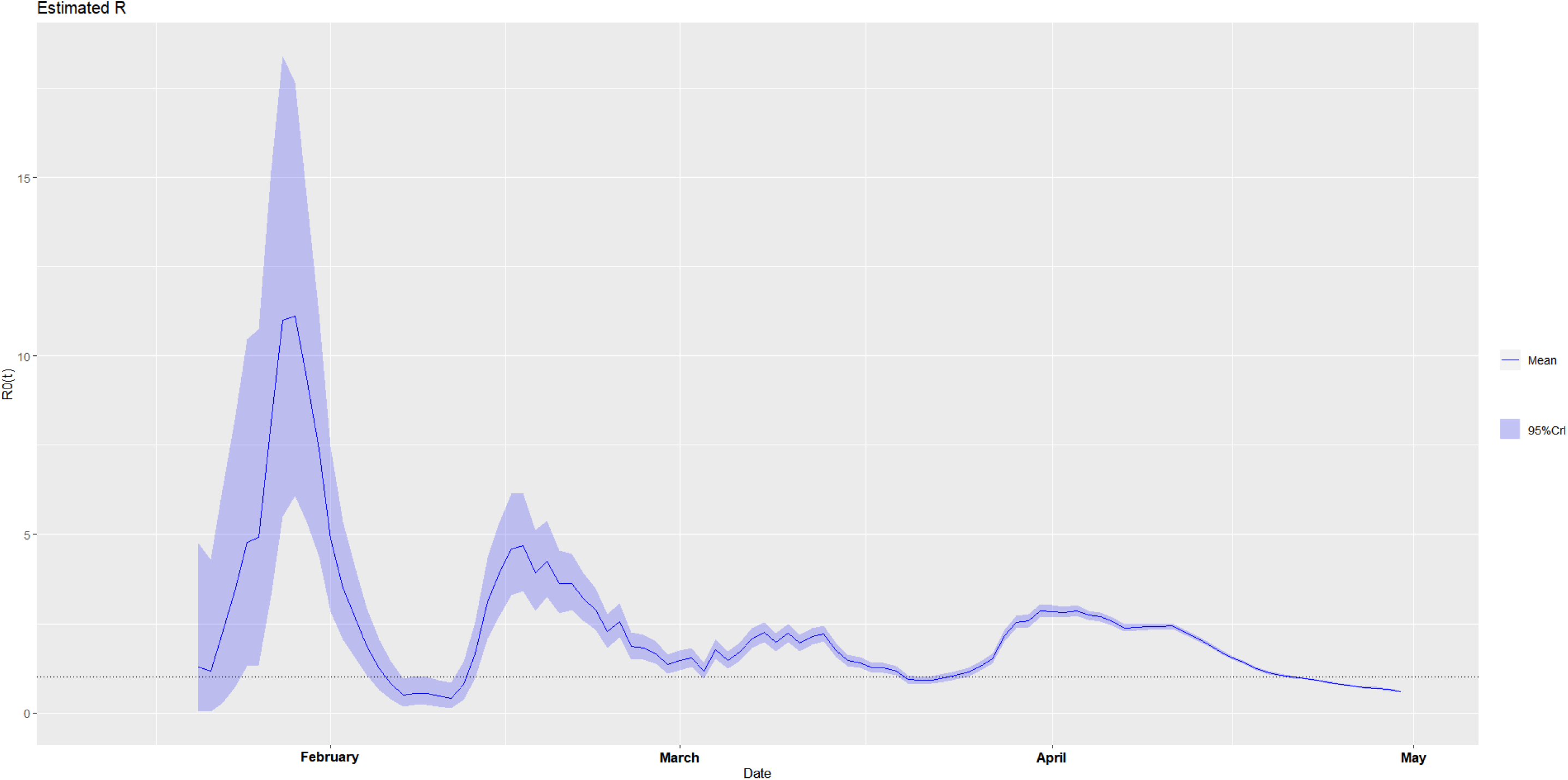
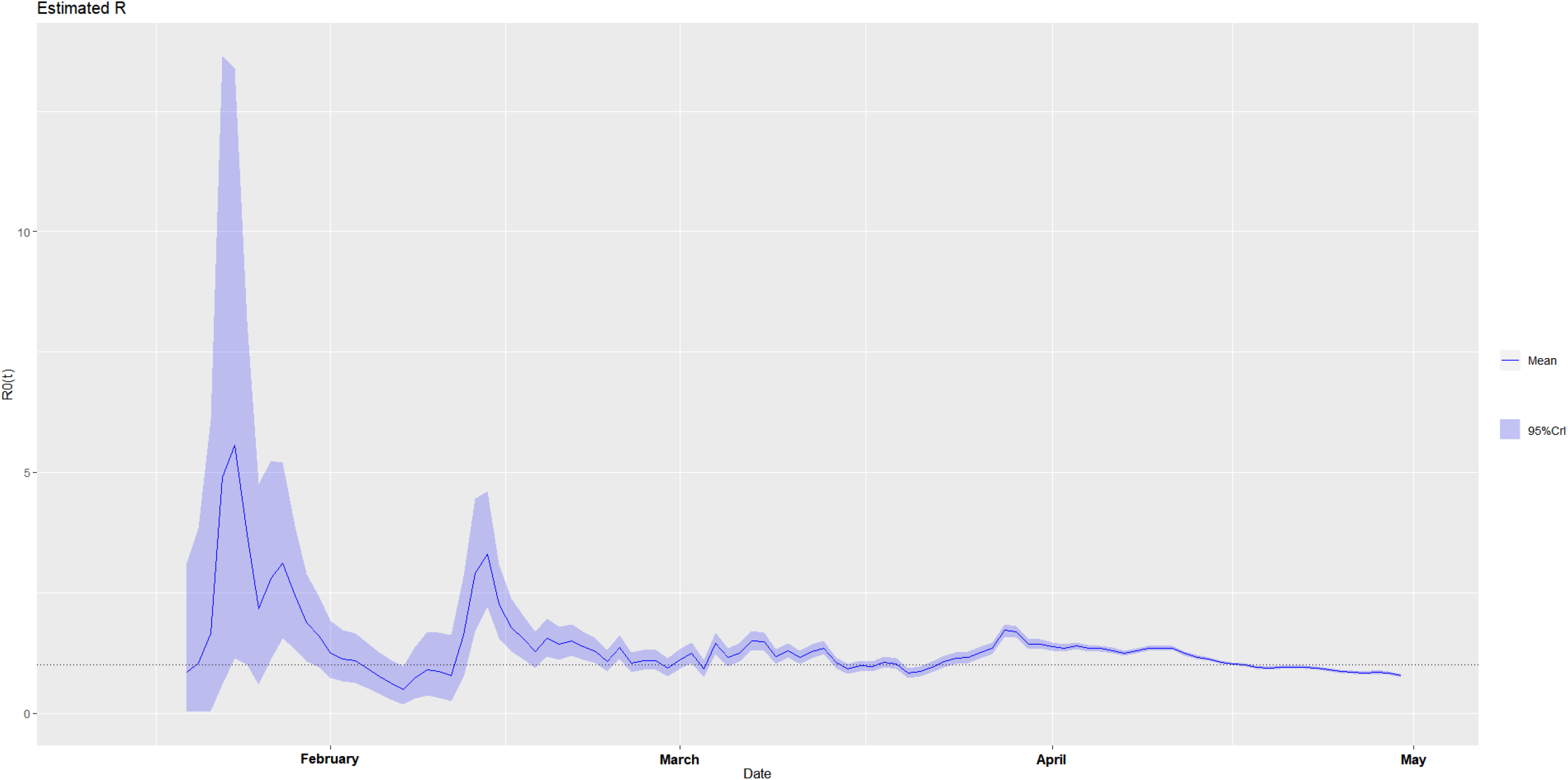
The estimated time-varying reproduction number during the ongoing pandemic of the coronavirus disease 2019 (COVID-19) in Japan from January 13, 2020 to April 30, 2020 under two scenarios. **A. Scenario 1:** We specified the mean (SD) of the Gamma distribution of serial interval (SI) to be 8.4 (3.8) days to mimic the 2003 epidemic of the severe acute respiratory syndrome (SARS) in Hong Kong.^7,8^ **B. Scenario 2:** We specified the mean (SD) of the Gamma distribution of serial interval (SI) to be 2.6 (1.5) days to mimic the 1918 pandemic of influenza in Baltimore, Maryland.^7,8^

**Figure 4-1.**
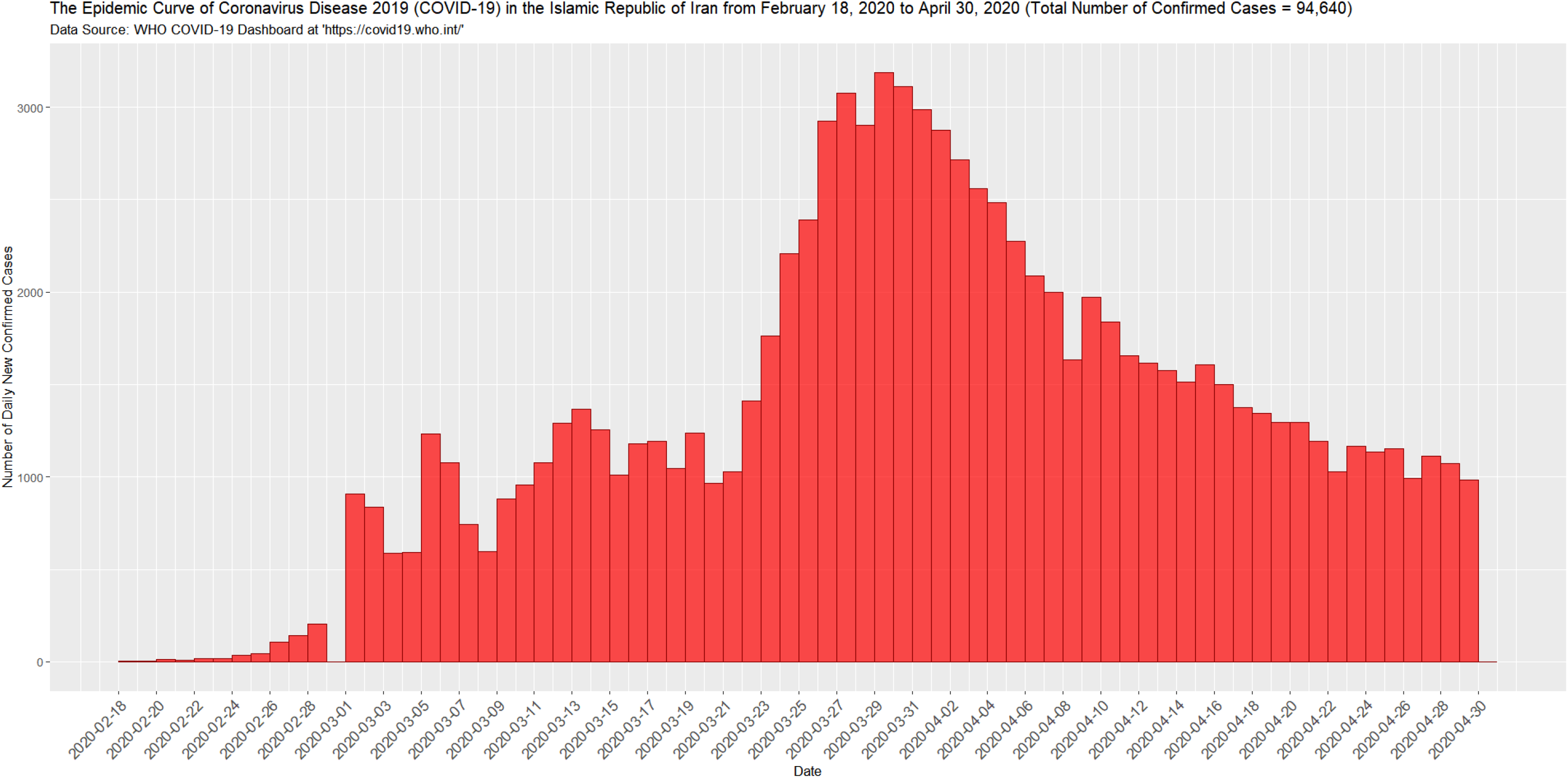
The epidemic curve of the coronavirus disease 2019 (COVID-19) in Iran from February 18, 2020 to April 30, 2020.

**Figure 4-2.**
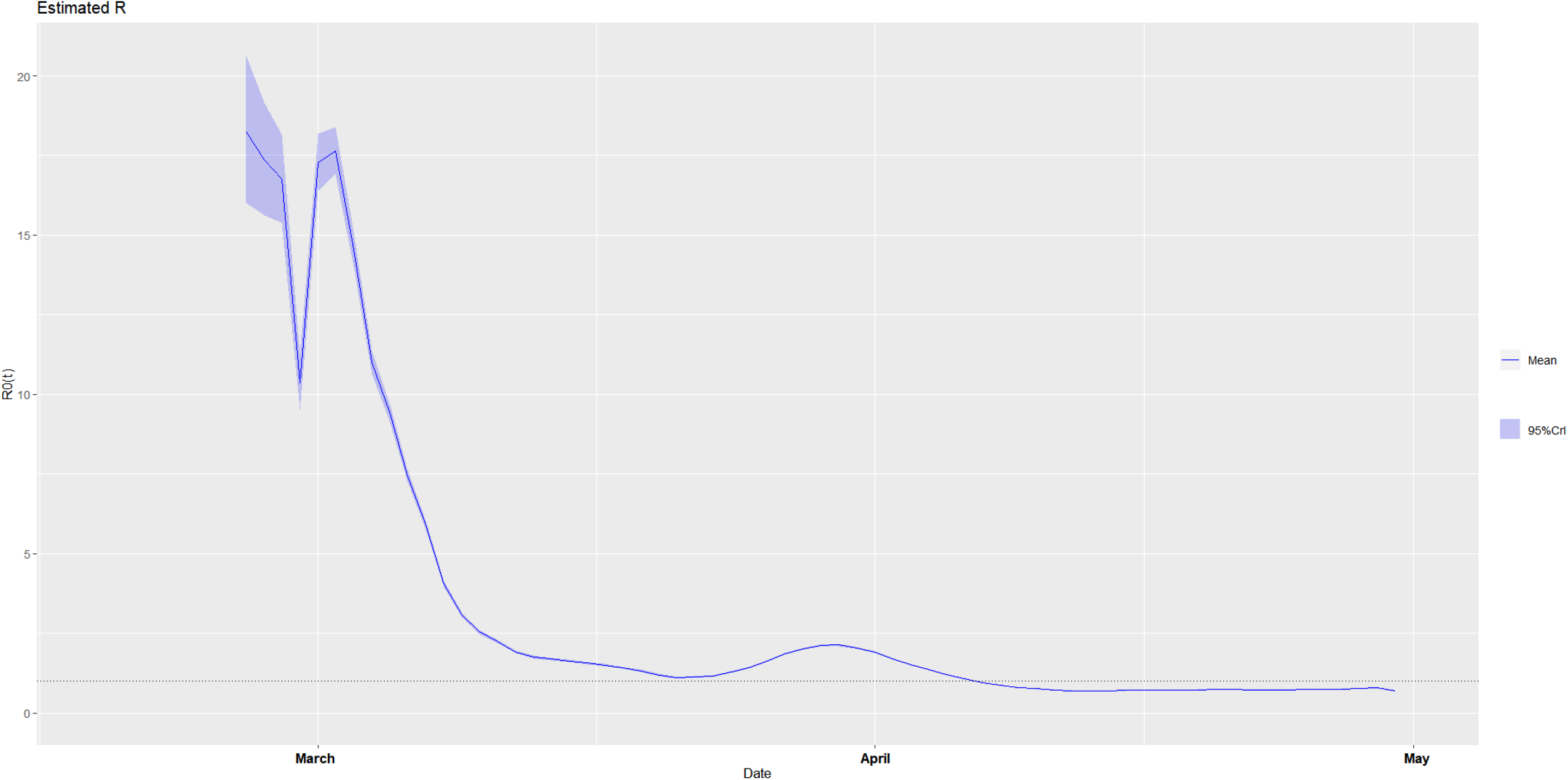
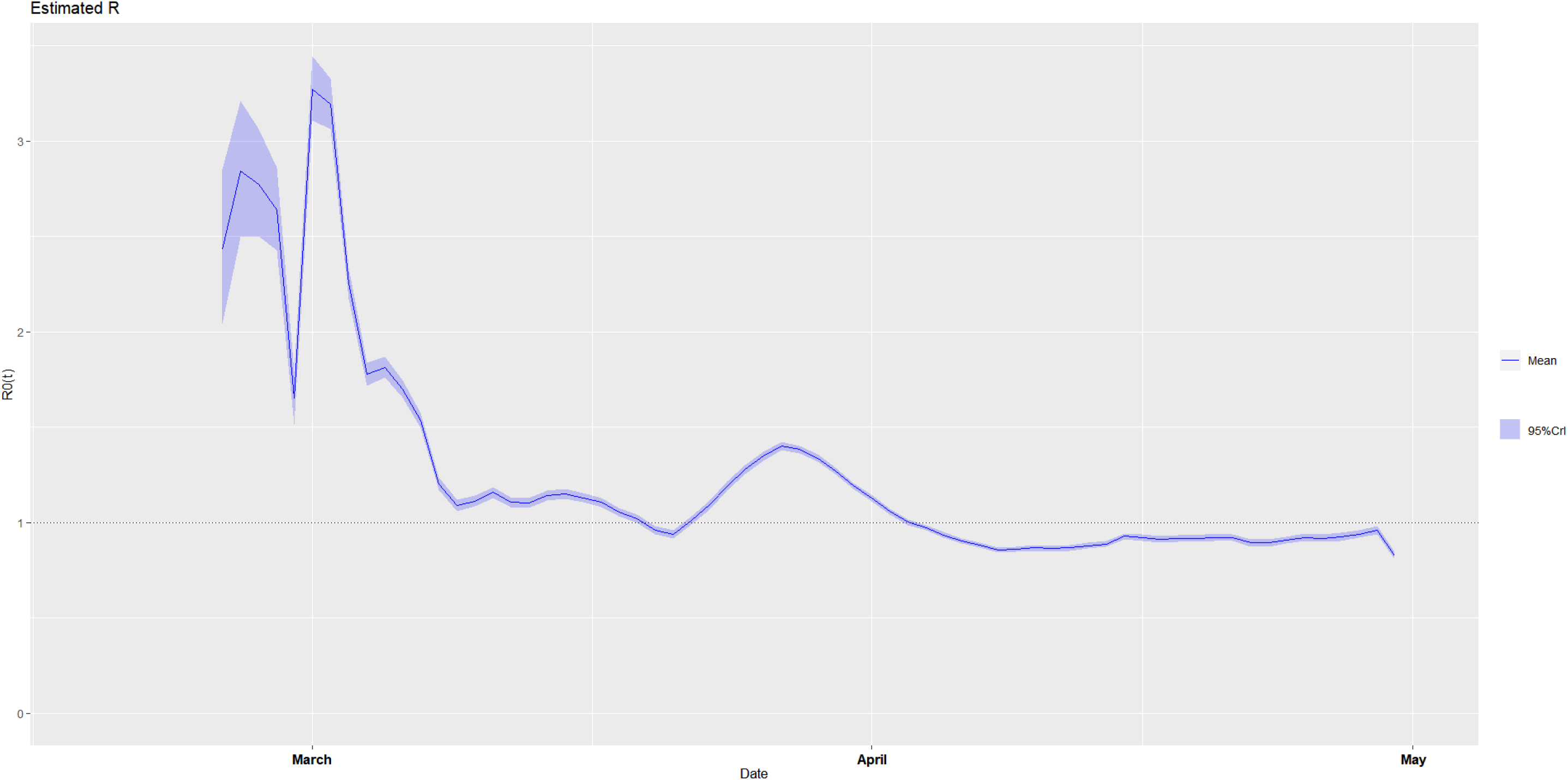
The estimated time-varying reproduction number during the ongoing pandemic of the coronavirus disease 2019 (COVID-19) in Iran from February 18, 2020 to April 30, 2020 under two scenarios. **A. Scenario 1:** We specified the mean (SD) of the Gamma distribution of serial interval (SI) to be 8.4 (3.8) days to mimic the 2003 epidemic of the severe acute respiratory syndrome (SARS) in Hong Kong.^7,8^ **B. Scenario 2:** We specified the mean (SD) of the Gamma distribution of serial interval (SI) to be 2.6 (1.5) days to mimic the 1918 pandemic of influenza in Baltimore, Maryland.^7,8^

**Figure 5-1.**
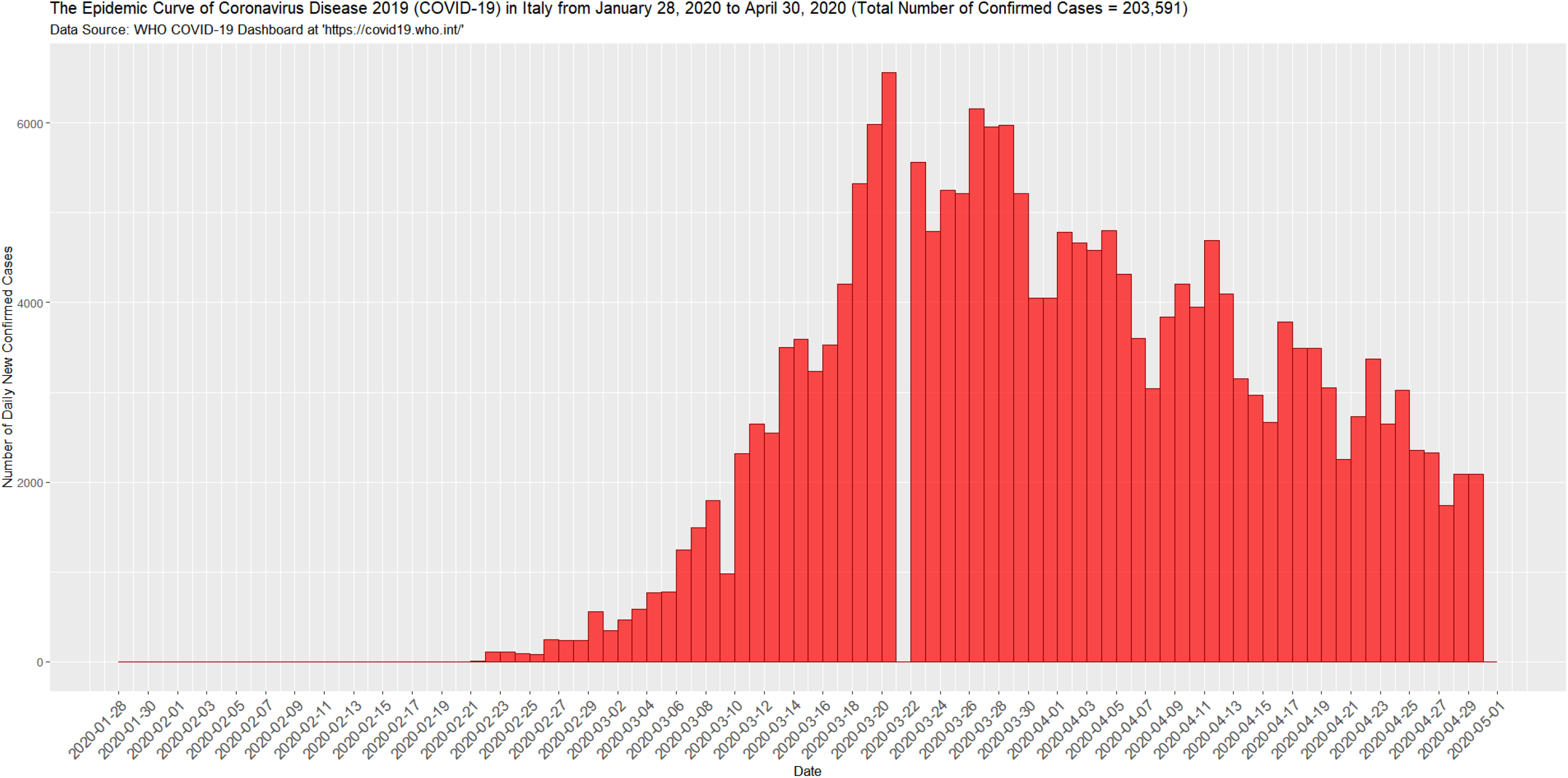
The epidemic curve of the coronavirus disease 2019 (COVID-19) in Italy from January 28, 2020 to April 30, 2020.

**Figure 5-2.**
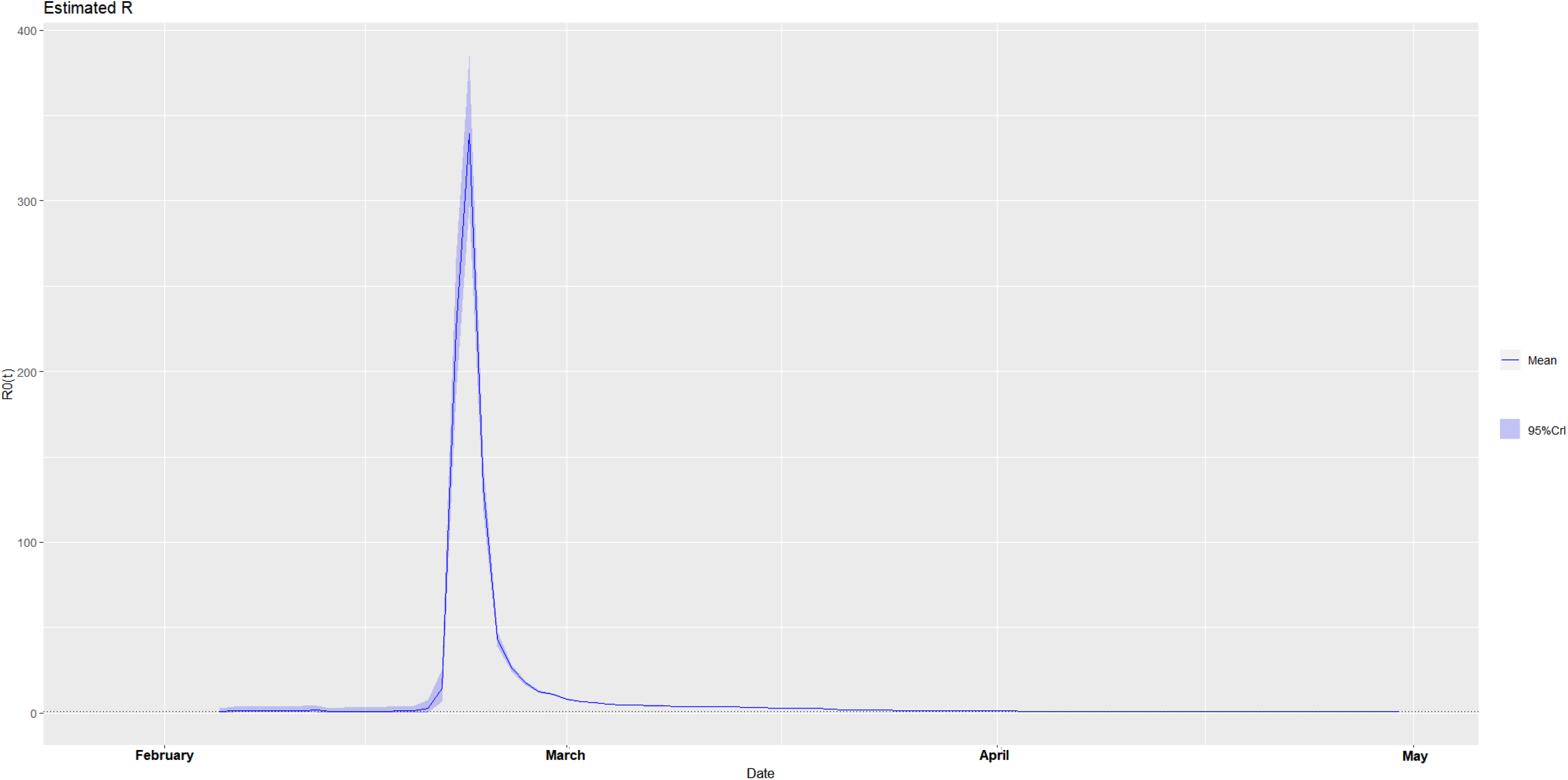
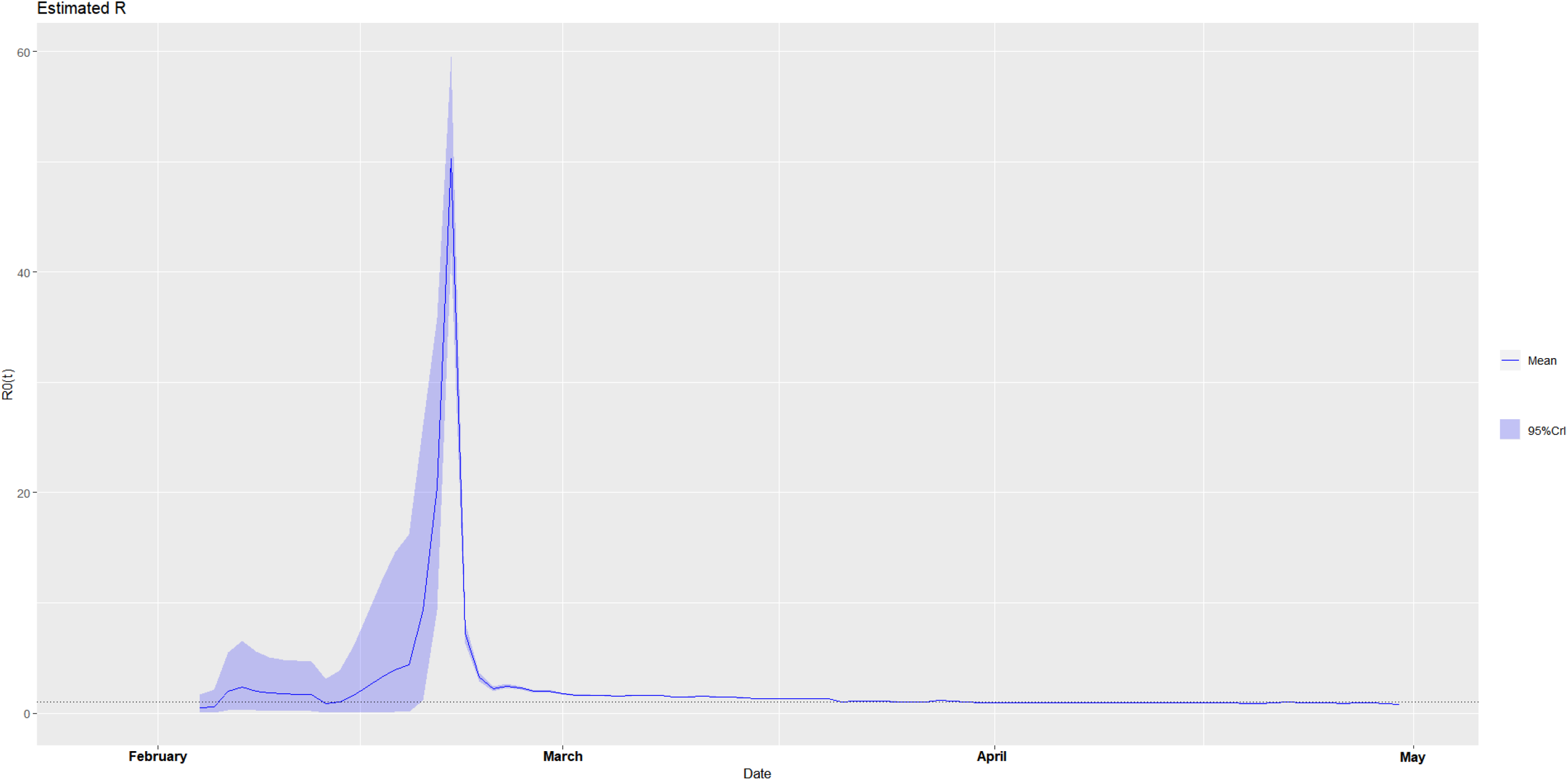
The estimated time-varying reproduction number during the ongoing pandemic of the coronavirus disease 2019 (COVID-19) in Italy from January 28, 2020 to April 30, 2020 under two scenarios. **A. Scenario 1:** We specified the mean (SD) of the Gamma distribution of serial interval (SI) to be 8.4 (3.8) days to mimic the 2003 epidemic of the severe acute respiratory syndrome (SARS) in Hong Kong.^7,8^ **B. Scenario 2:** We specified the mean (SD) of the Gamma distribution of serial interval (SI) to be 2.6 (1.5) days to mimic the 1918 pandemic of influenza in Baltimore, Maryland.^7,8^

**Figure 6-1.**
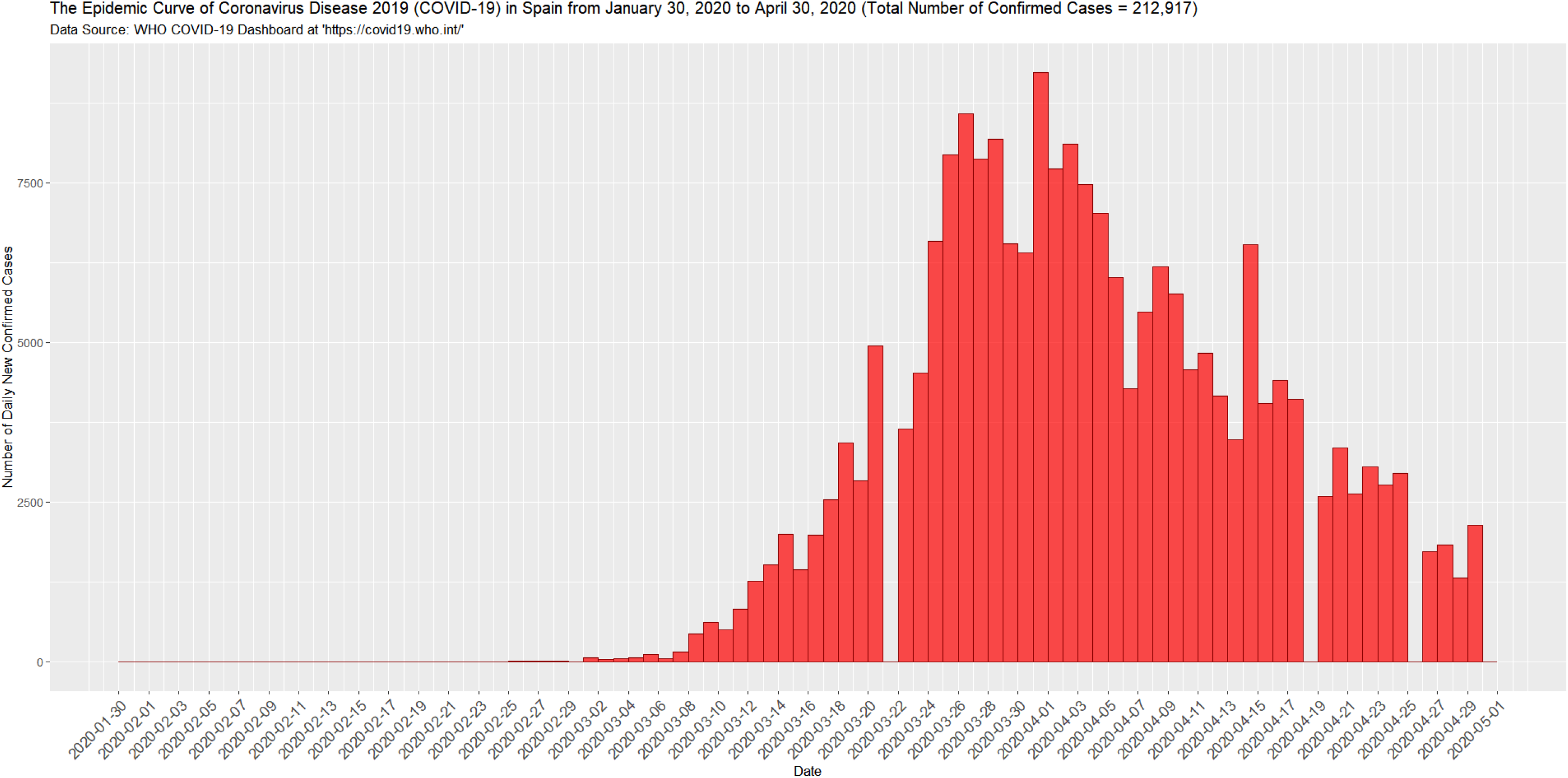
The epidemic curve of the coronavirus disease 2019 (COVID-19) in Spain from January 30, 2020 to April 30, 2020.

**Figure 6-2.**
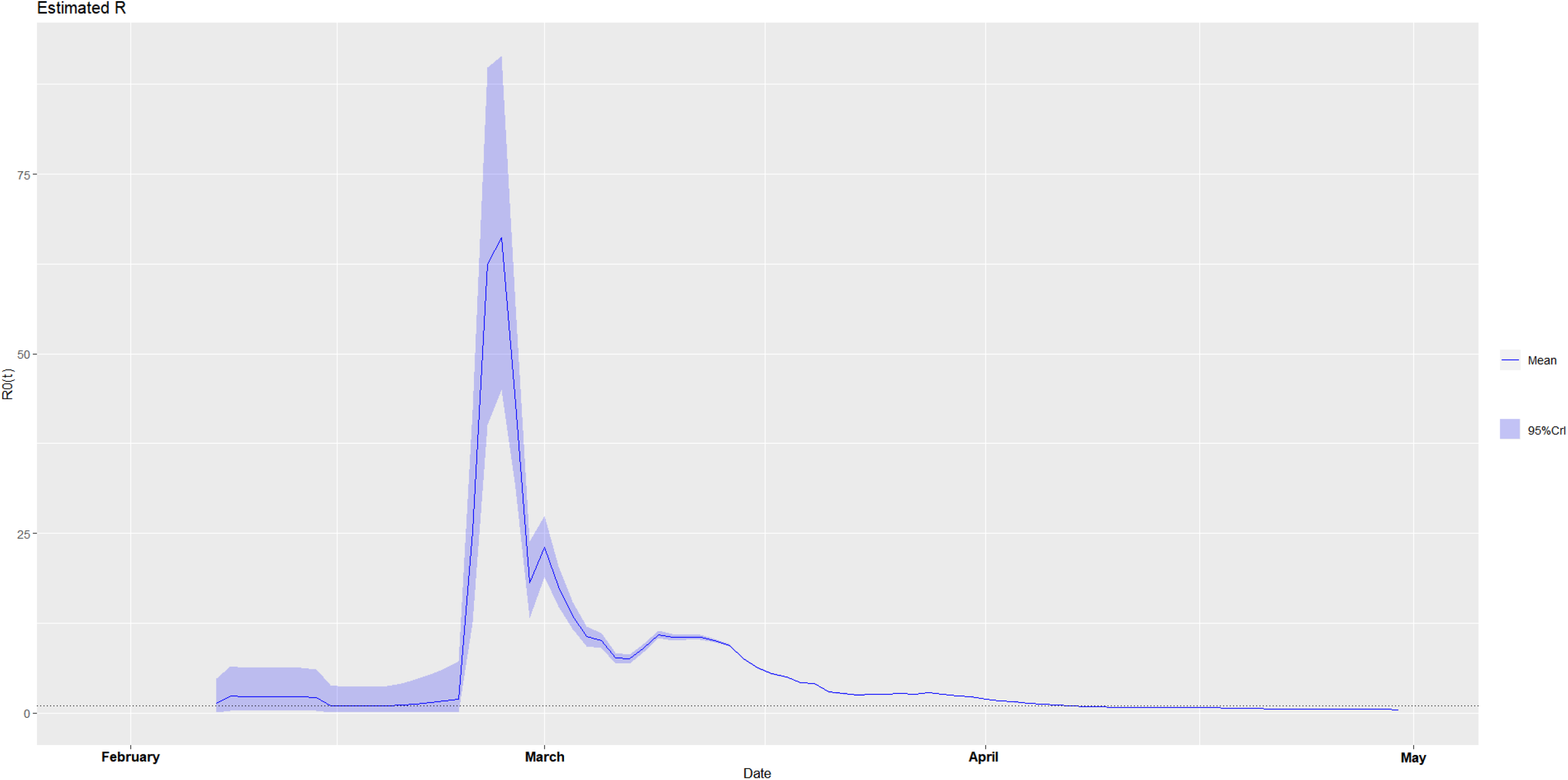
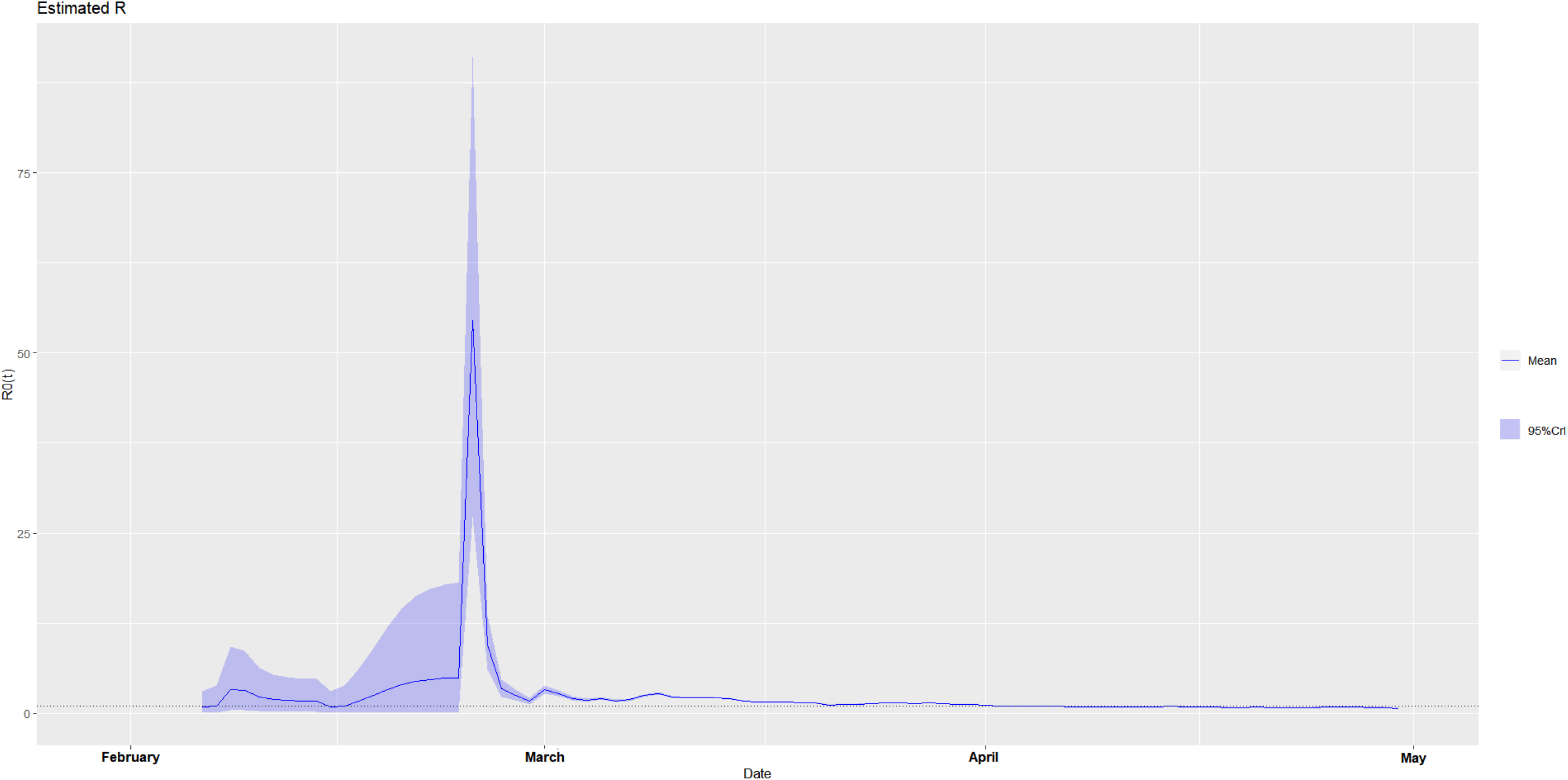
The estimated time-varying reproduction number during the ongoing pandemic of the coronavirus disease 2019 (COVID-19) in Spain from January 30, 2020 to April 30, 2020 under two scenarios. **A. Scenario 1:** We specified the mean (SD) of the Gamma distribution of serial interval (SI) to be 8.4 (3.8) days to mimic the 2003 epidemic of the severe acute respiratory syndrome (SARS) in Hong Kong.^7,8^ **B. Scenario 2:** We specified the mean (SD) of the Gamma distribution of serial interval (SI) to be 2.6 (1.5) days to mimic the 1918 pandemic of influenza in Baltimore, Maryland.^7,8^

**Figure 7-1.**
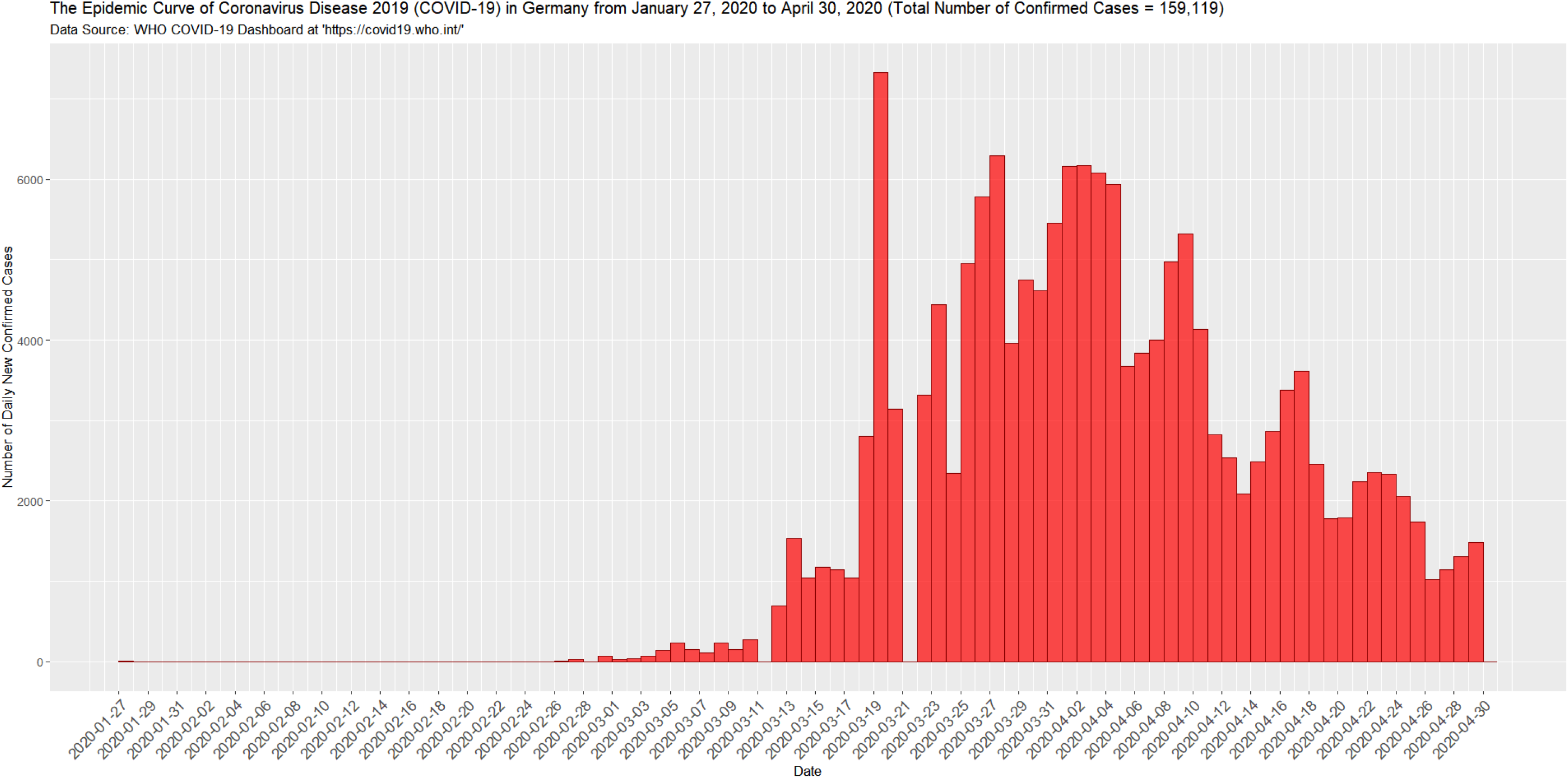
The epidemic curve of the coronavirus disease 2019 (COVID-19) in Germany from January 27, 2020 to April 30, 2020.

**Figure 7-2.**
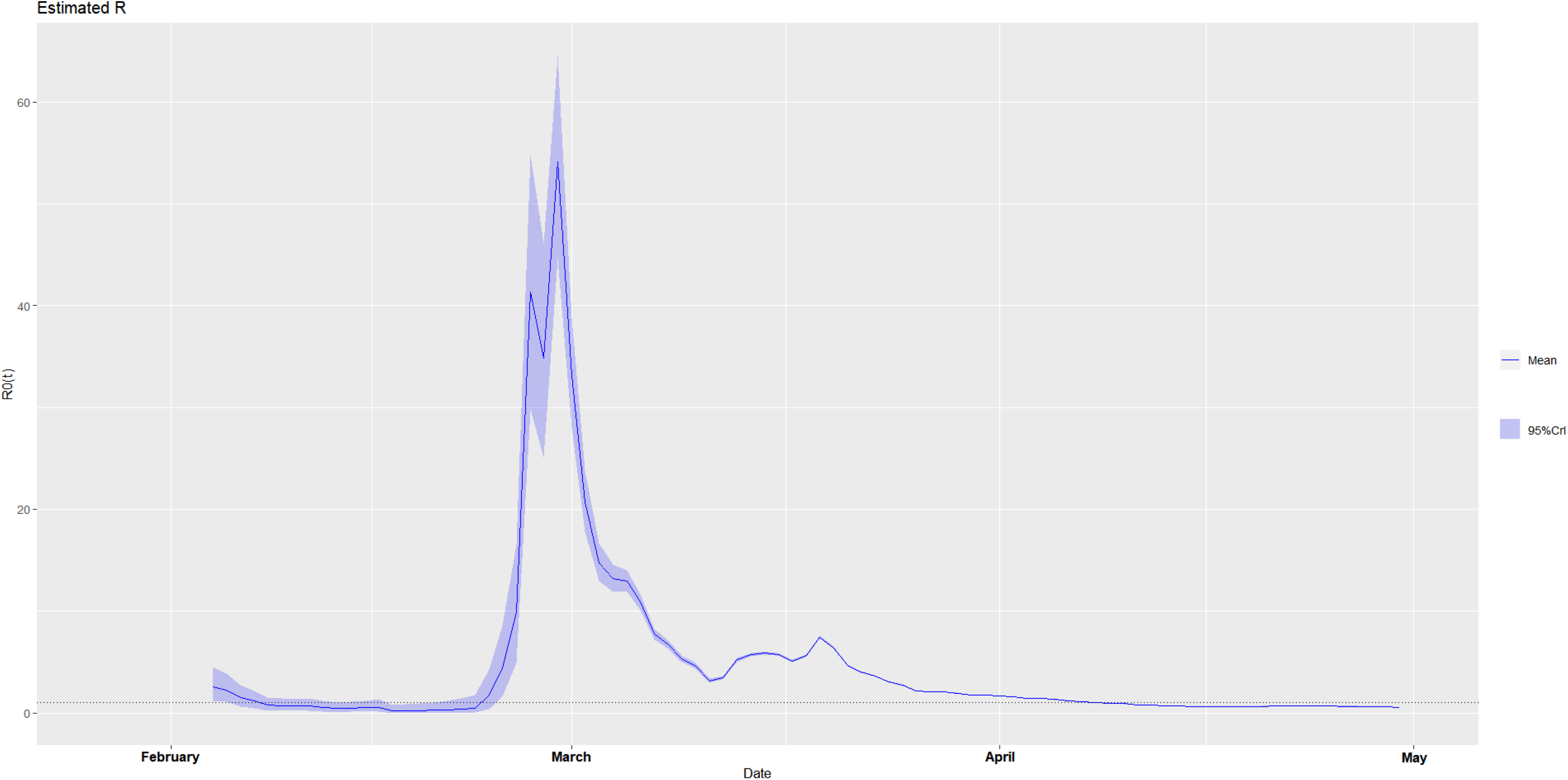
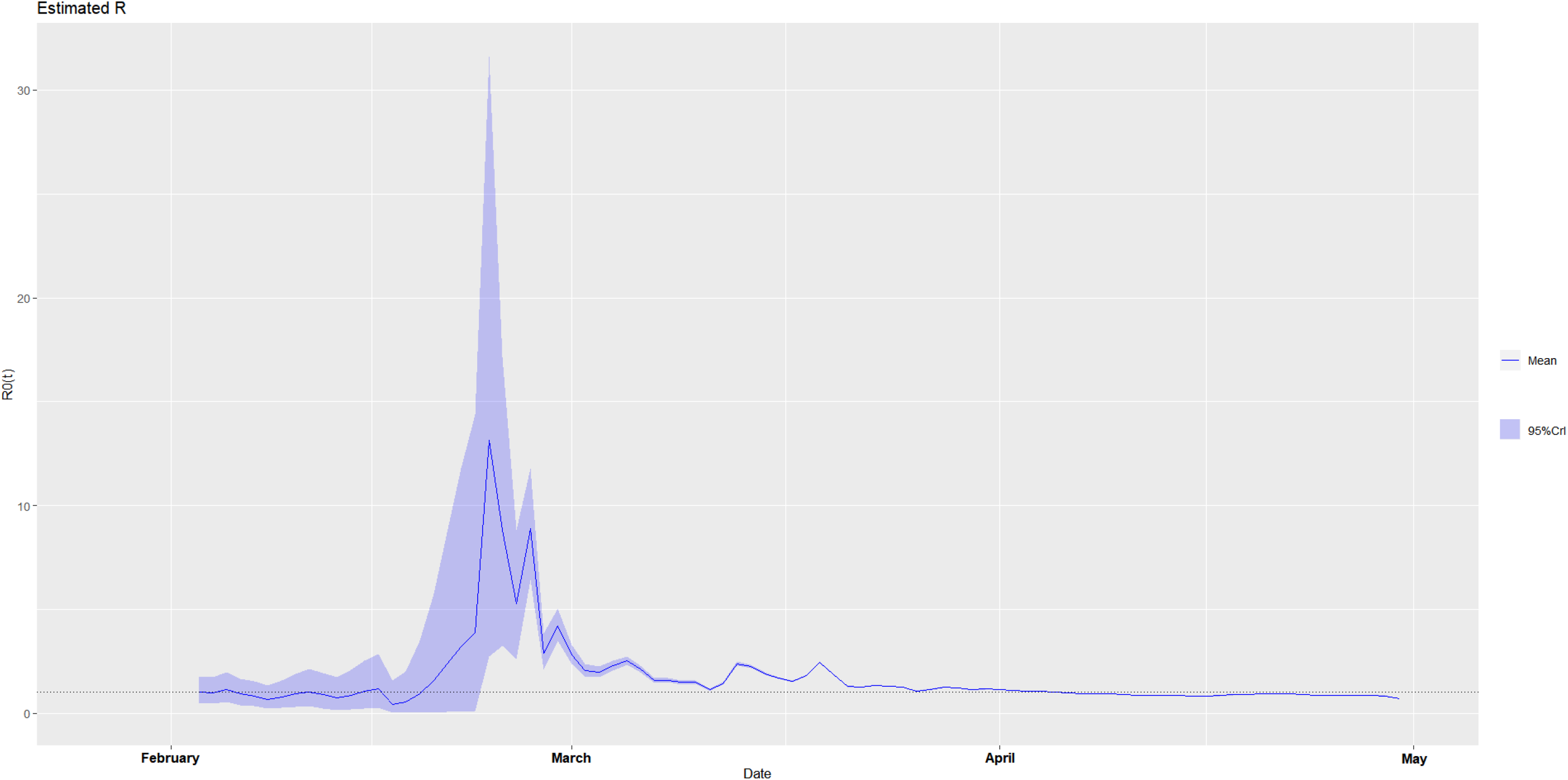
The estimated time-varying reproduction number during the ongoing pandemic of the coronavirus disease 2019 (COVID-19) in Germany from January 27, 2020 to April 30, 2020 under two scenarios. **A. Scenario 1:** We specified the mean (SD) of the Gamma distribution of serial interval (SI) to be 8.4 (3.8) days to mimic the 2003 epidemic of the severe acute respiratory syndrome (SARS) in Hong Kong.^7,8^ **B. Scenario 2:** We specified the mean (SD) of the Gamma distribution of serial interval (SI) to be 2.6 (1.5) days to mimic the 1918 pandemic of influenza in Baltimore, Maryland.^7,8^

**Figure 8-1.**
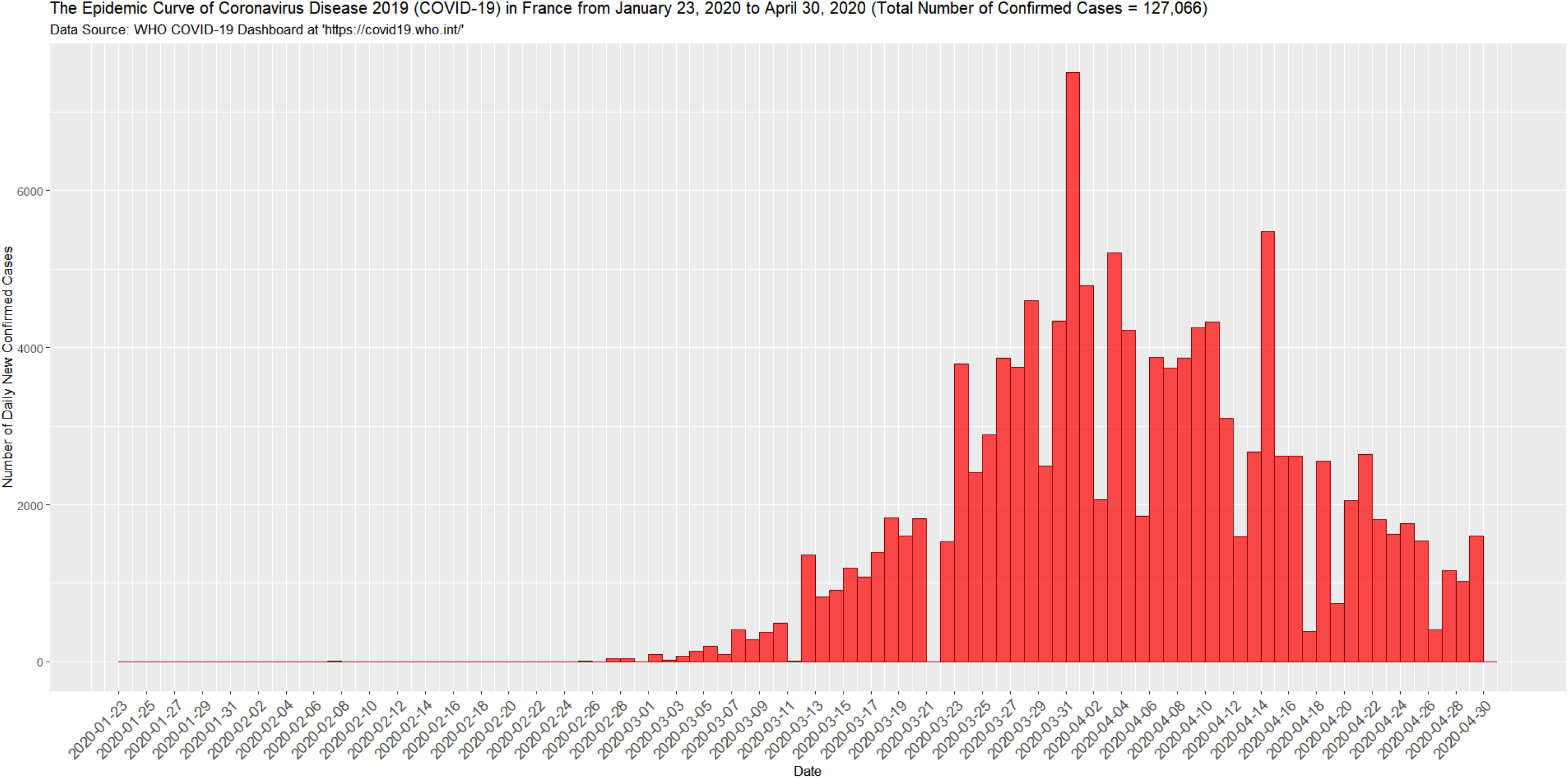
The epidemic curve of the coronavirus disease 2019 (COVID-19) in France from January 23, 2020 to April 30, 2020.

**Figure 8-2.**
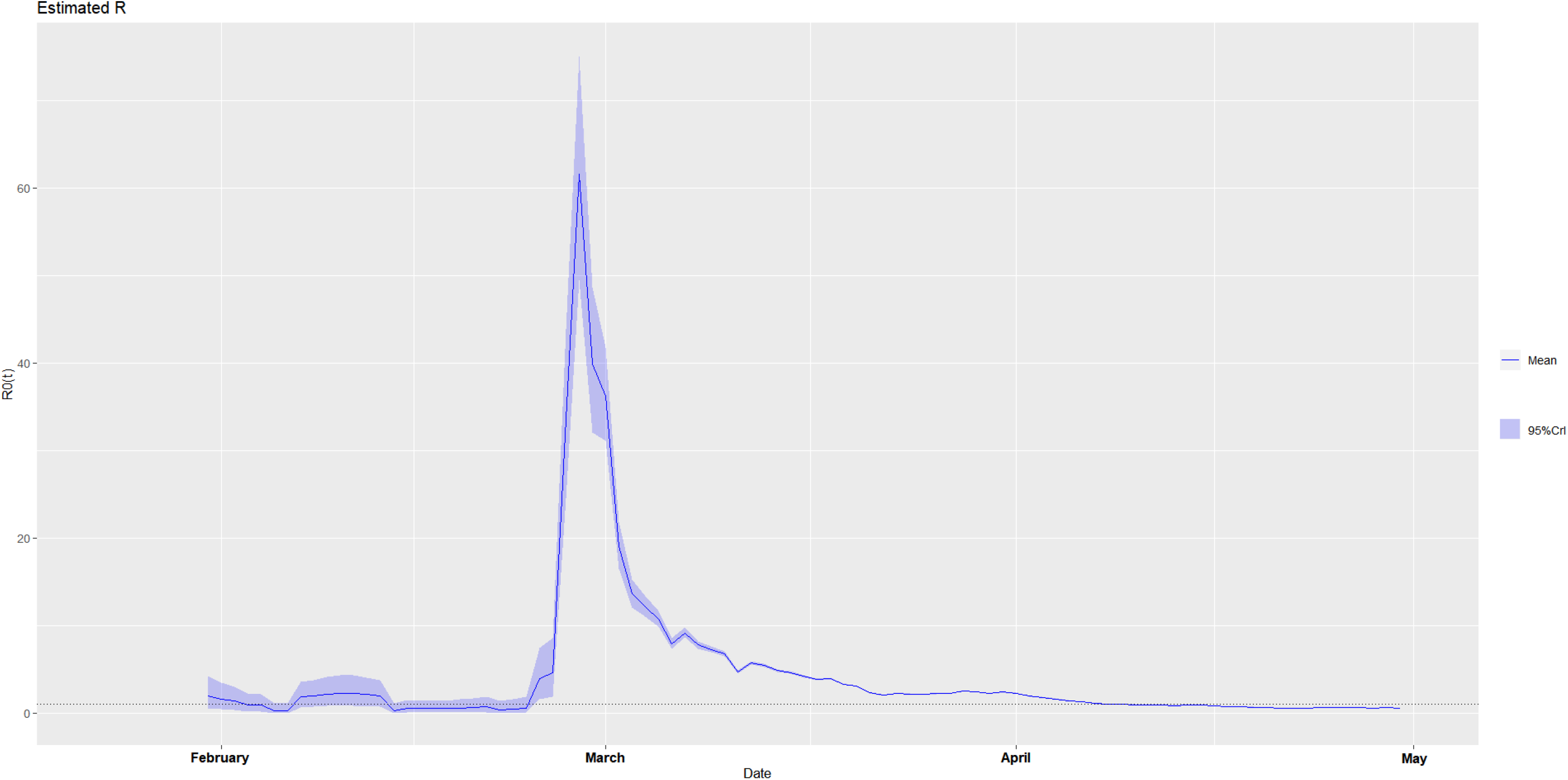
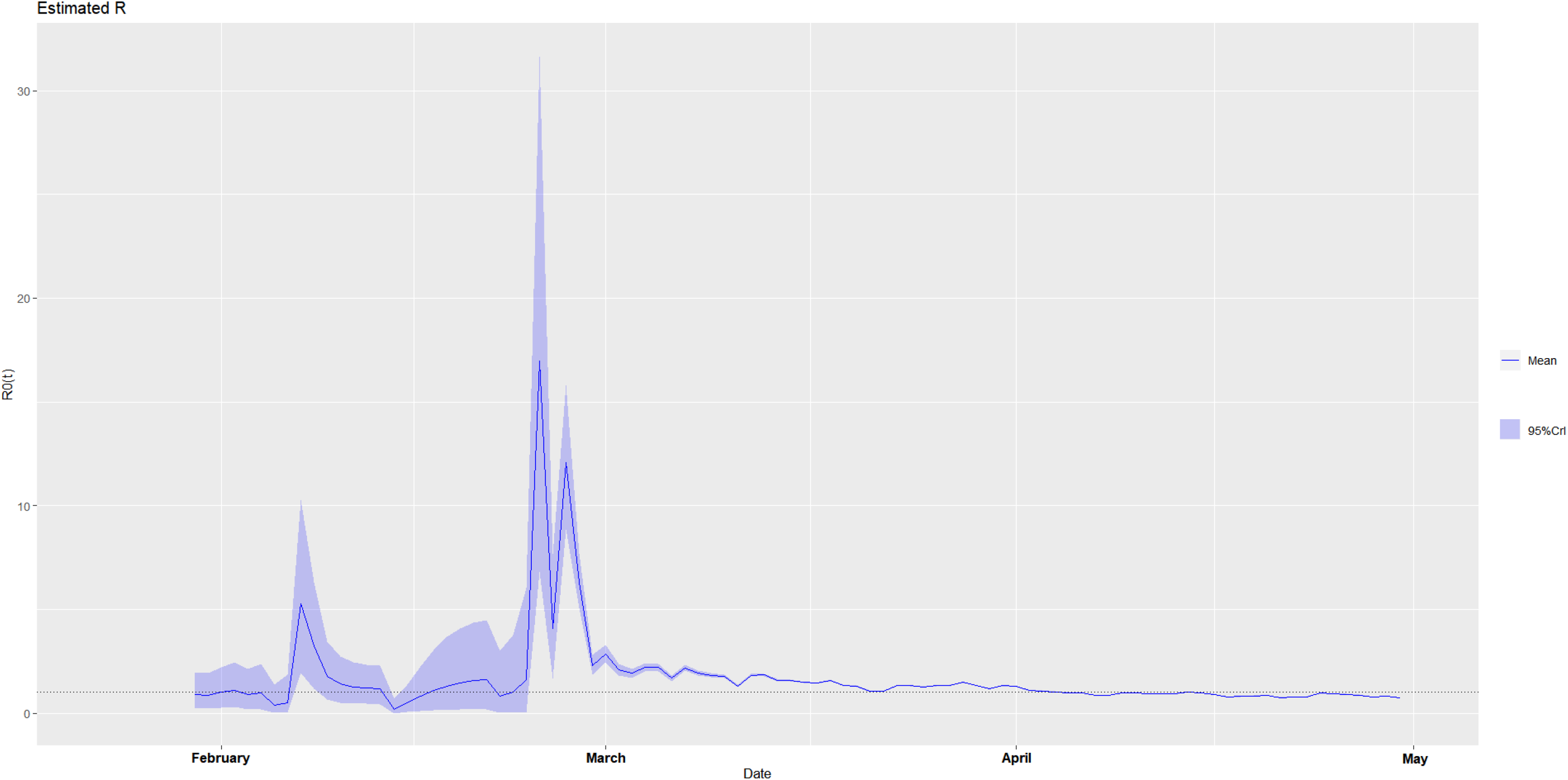
The estimated time-varying reproduction number during the ongoing pandemic of the coronavirus disease 2019 (COVID-19) in France from January 23, 2020 to April 30, 2020 under two scenarios. **A. Scenario 1:** We specified the mean (SD) of the Gamma distribution of serial interval (SI) to be 8.4 (3.8) days to mimic the 2003 epidemic of the severe acute respiratory syndrome (SARS) in Hong Kong.^7,8^ **B. Scenario 2:** We specified the mean (SD) of the Gamma distribution of serial interval (SI) to be 2.6 (1.5) days to mimic the 1918 pandemic of influenza in Baltimore, Maryland.^7,8^

**Figure 9-1.**
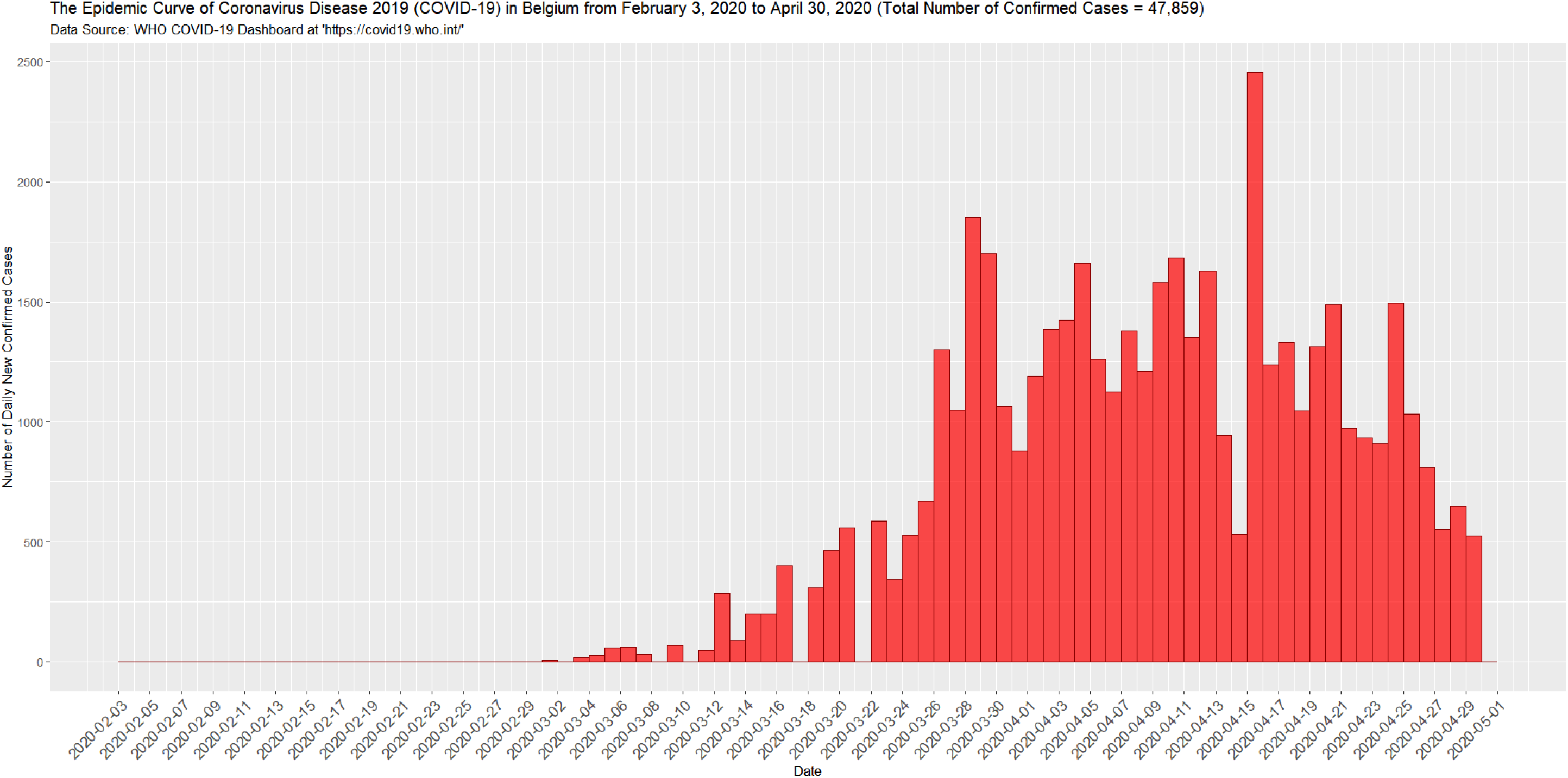
The epidemic curve of the coronavirus disease 2019 (COVID-19) in Belgium from February 3, 2020 to April 30, 2020.

**Figure 9-2.**
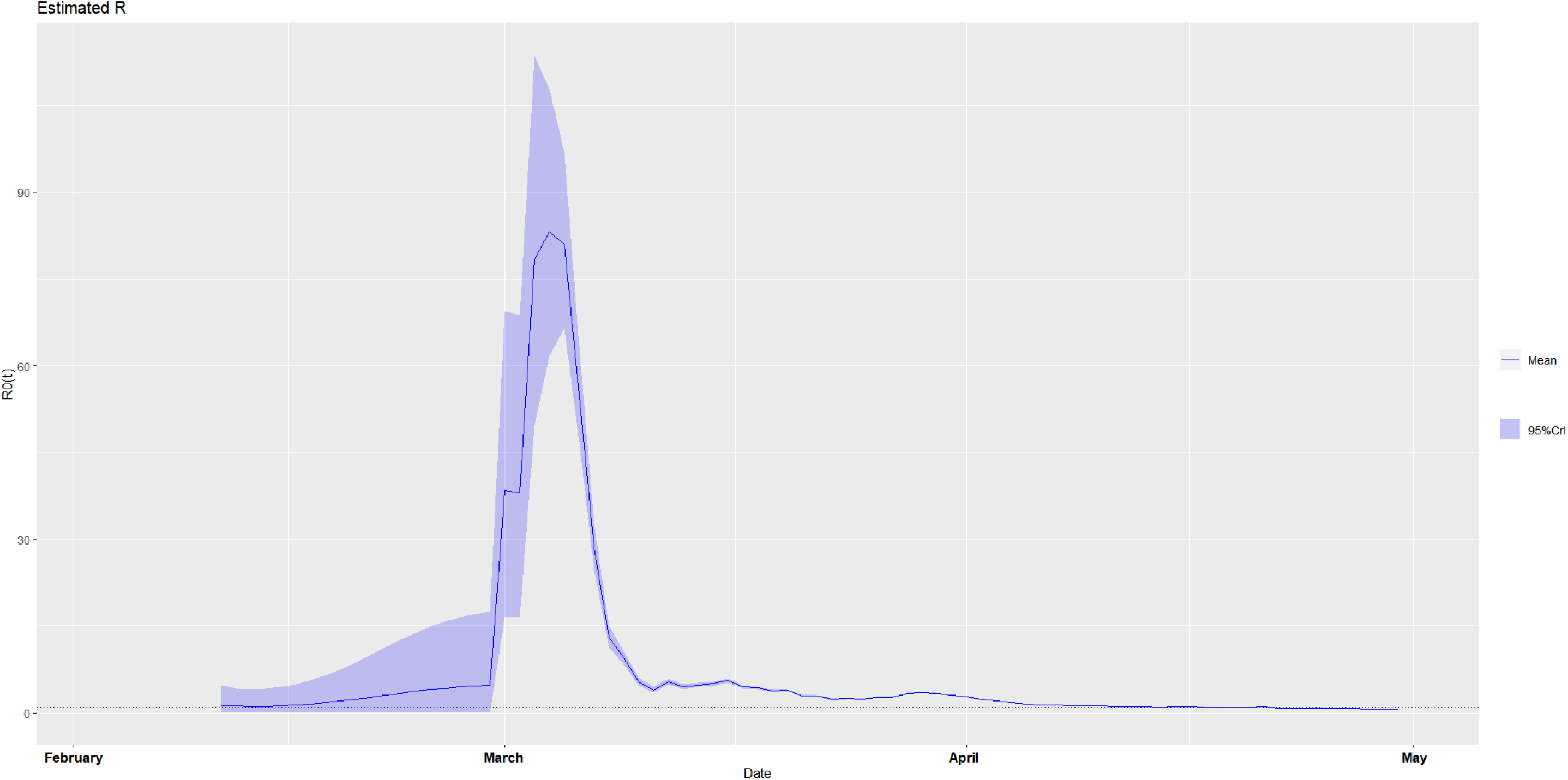
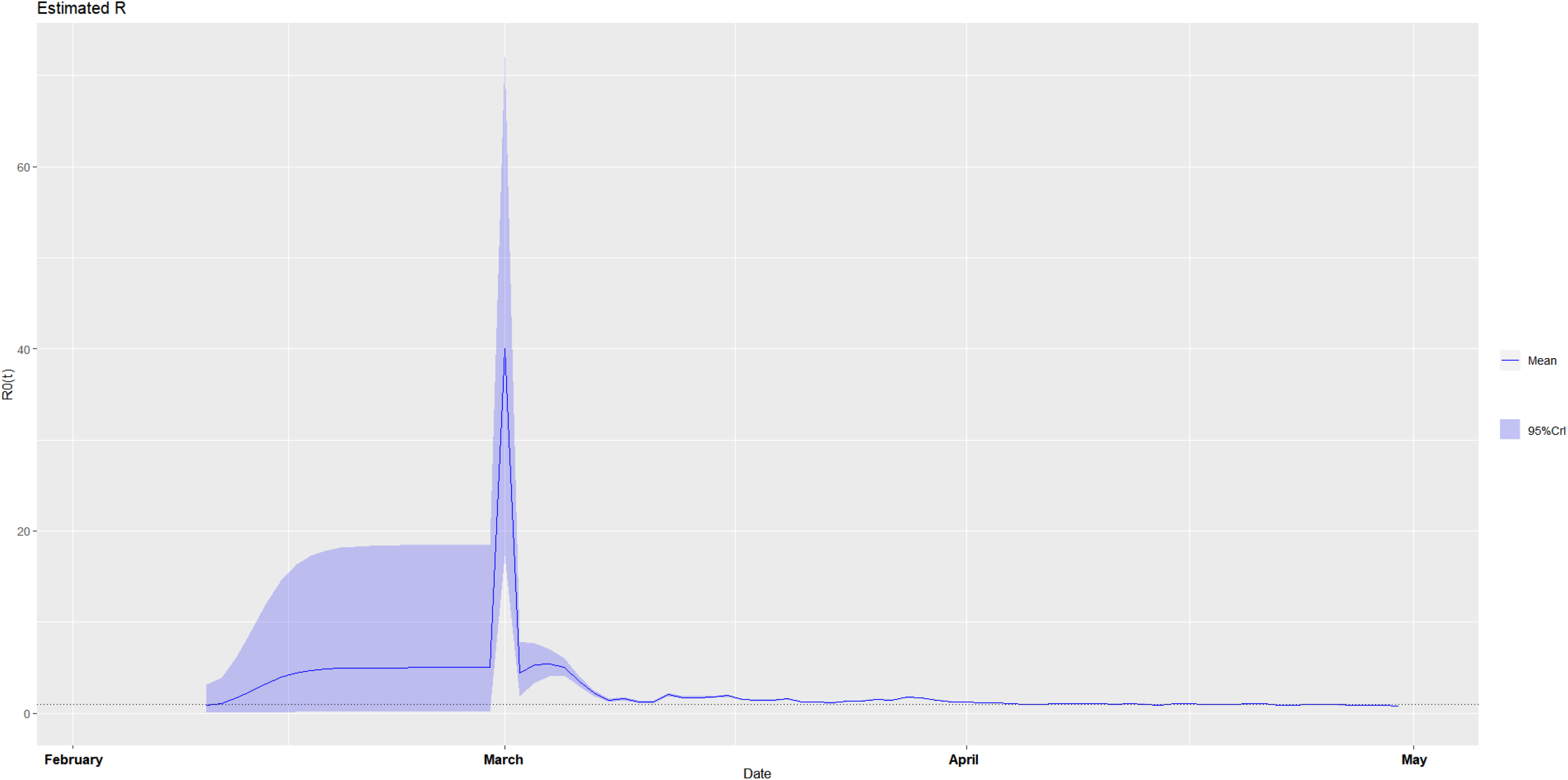
The estimated time-varying reproduction number during the ongoing pandemic of the coronavirus disease 2019 (COVID-19) in Belgium from February 3, 2020 to April 30, 2020 under two scenarios. **A. Scenario 1:** We specified the mean (SD) of the Gamma distribution of serial interval (SI) to be 8.4 (3.8) days to mimic the 2003 epidemic of the severe acute respiratory syndrome (SARS) in Hong Kong.^7,8^ **B. Scenario 2:** We specified the mean (SD) of the Gamma distribution of serial interval (SI) to be 2.6 (1.5) days to mimic the 1918 pandemic of influenza in Baltimore, Maryland.^7,8^

**Figure 10-1.**
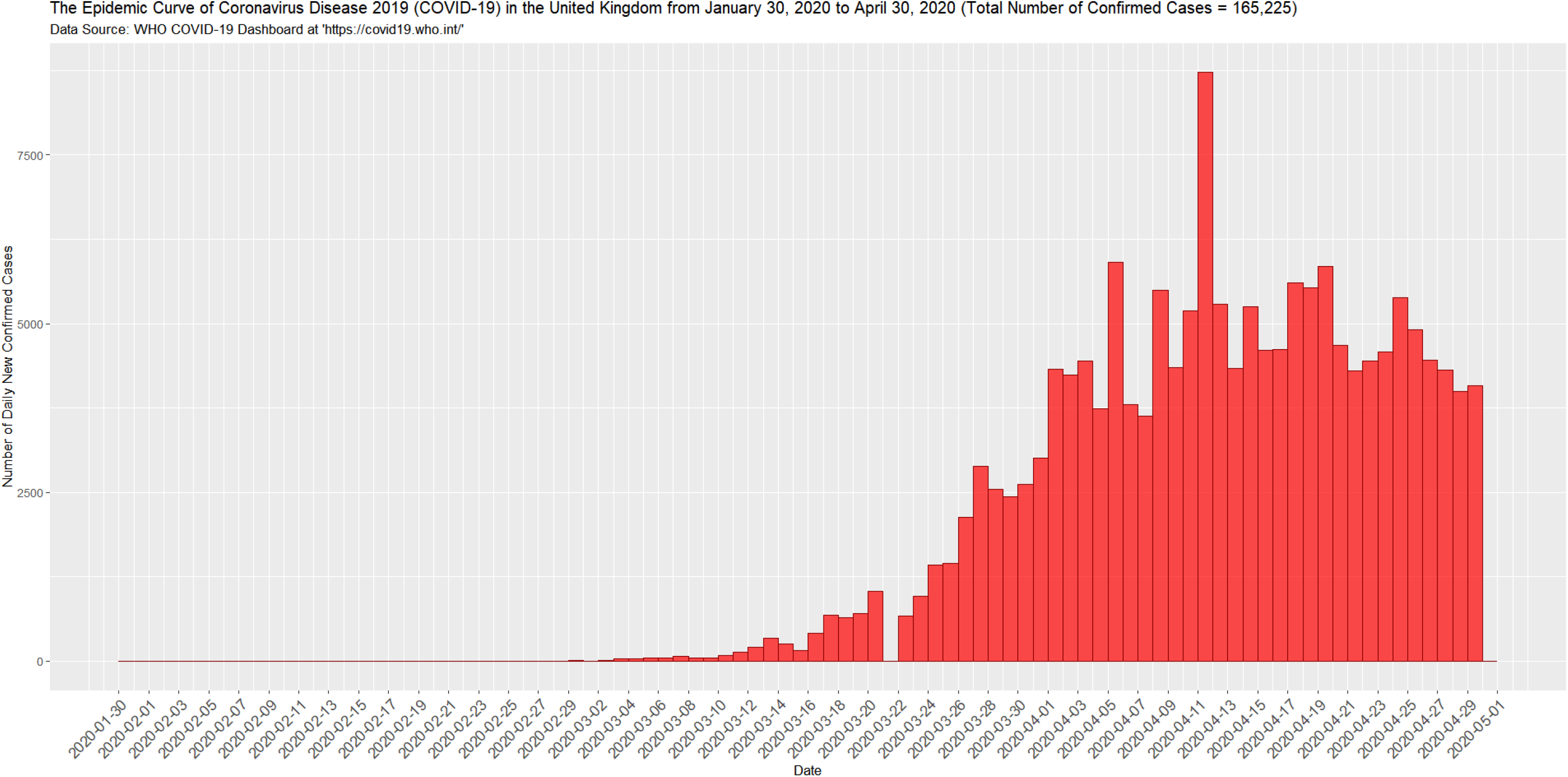
The epidemic curve of the coronavirus disease 2019 (COVID-19) in the United Kingdom from January 30, 2020 to April 30, 2020.

**Figure 10-2.**
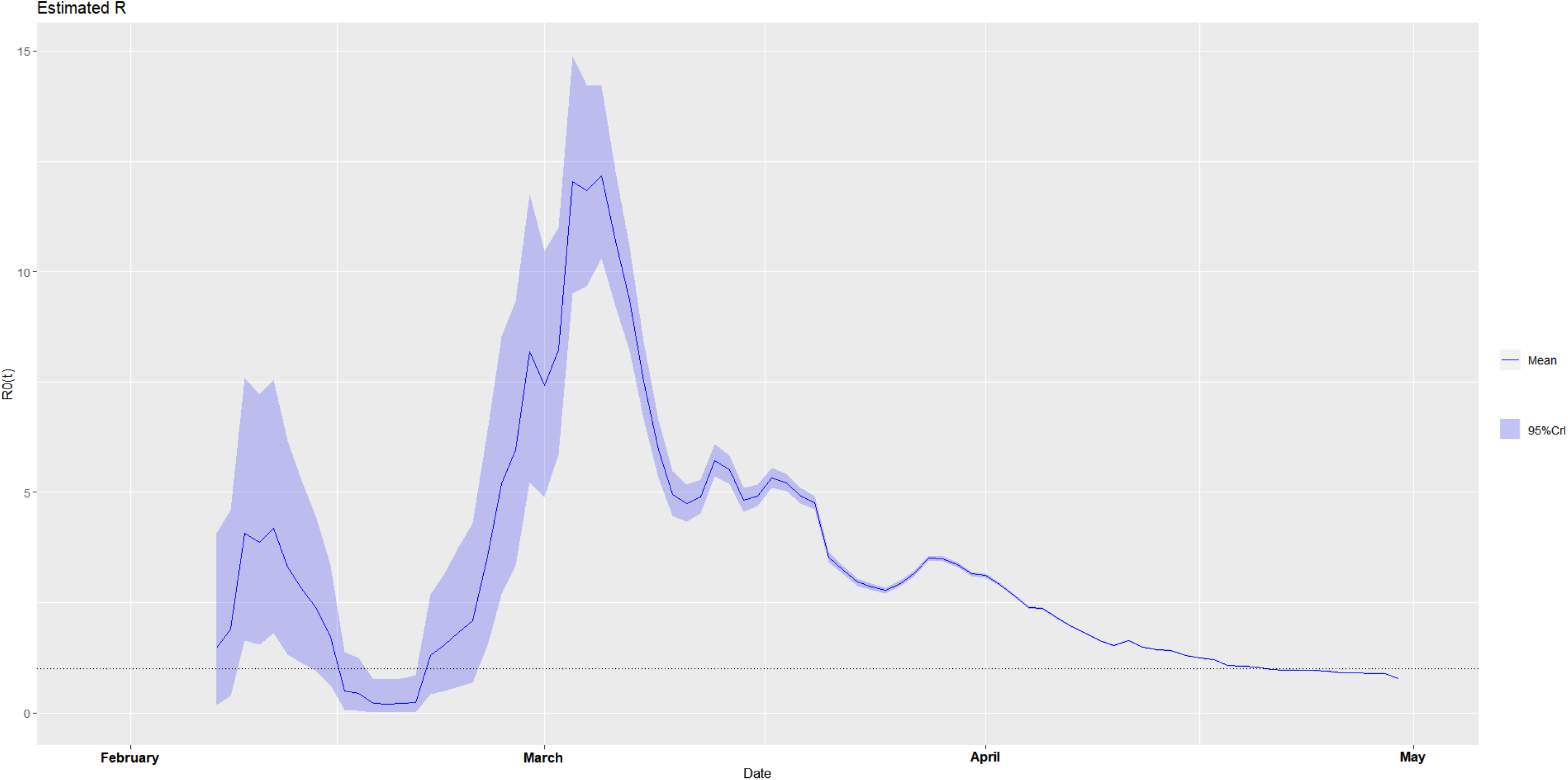
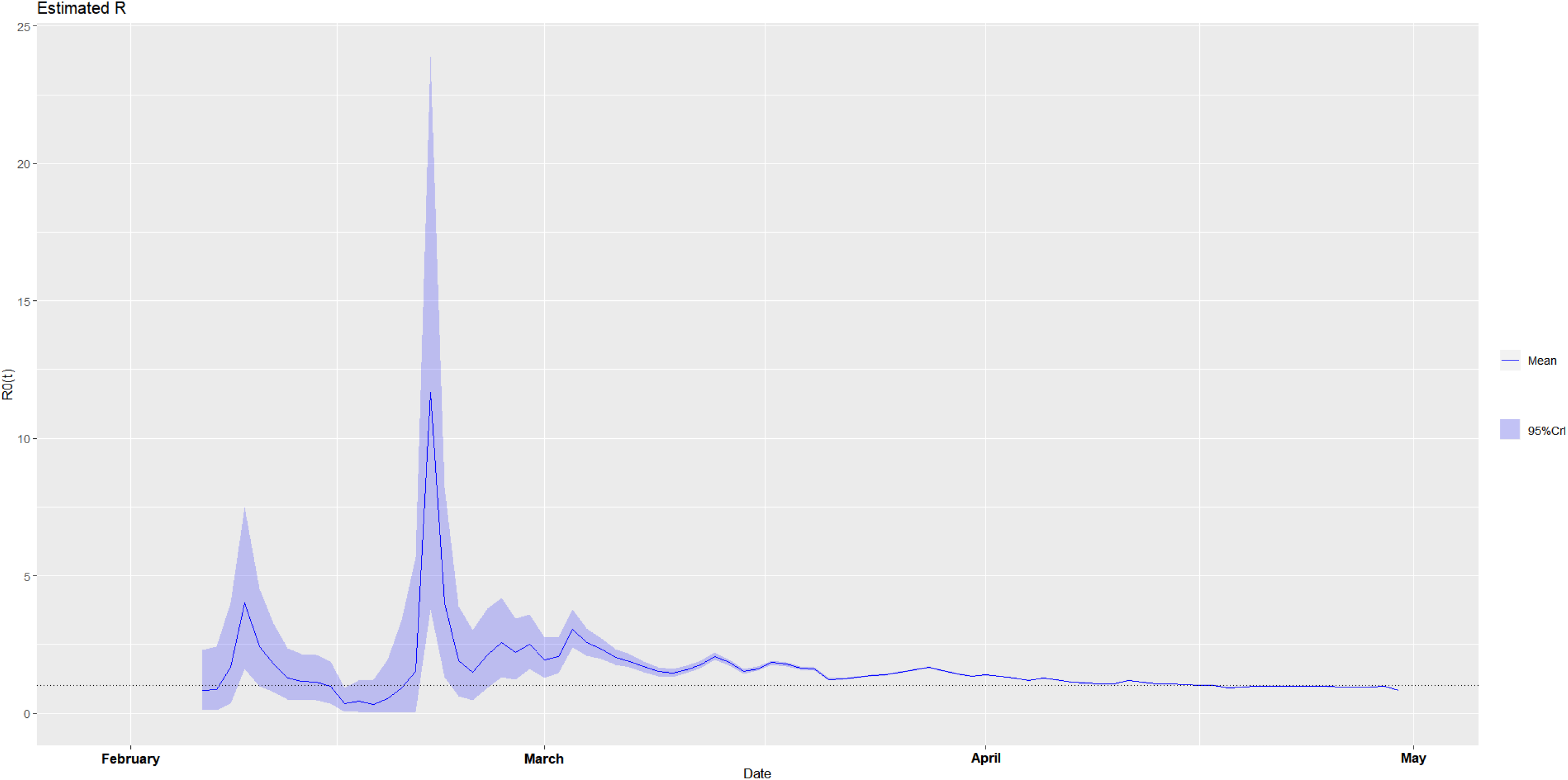
The estimated time-varying reproduction number during the ongoing pandemic of the coronavirus disease 2019 (COVID-19) in the United Kingdom from January 30, 2020 to April 30, 2020 under two scenarios. **A. Scenario 1:** We specified the mean (SD) of the Gamma distribution of serial interval (SI) to be 8.4 (3.8) days to mimic the 2003 epidemic of the severe acute respiratory syndrome (SARS) in Hong Kong.^7,8^ **B. Scenario 2:** We specified the mean (SD) of the Gamma distribution of serial interval (SI) to be 2.6 (1.5) days to mimic the 1918 pandemic of influenza in Baltimore, Maryland.^7,8^

**Figure 11-1.**
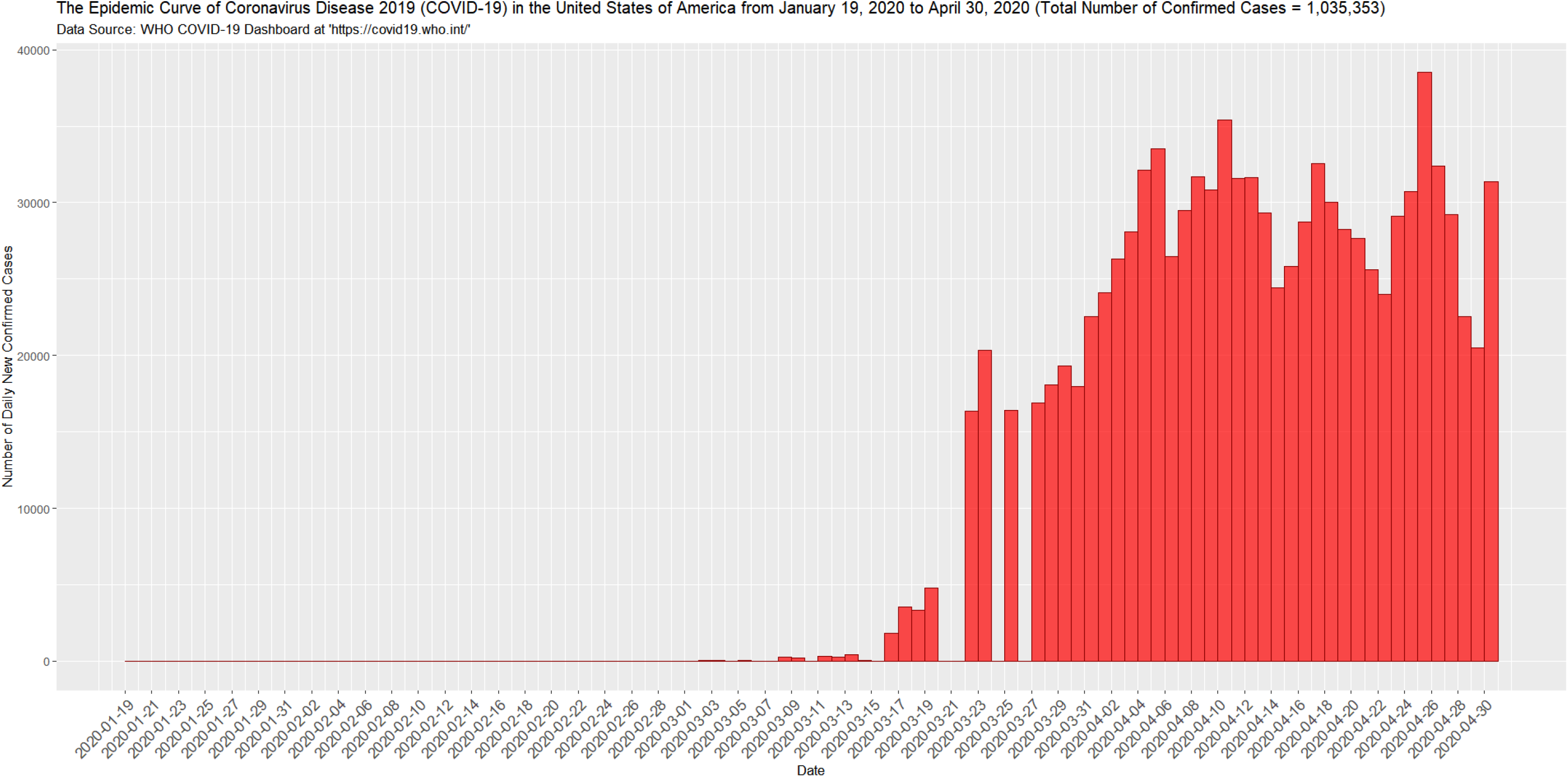
The epidemic curve of the coronavirus disease 2019 (COVID-19) in the United States of America from January 19, 2020 to April 30, 2020.

**Figure 11-2.**
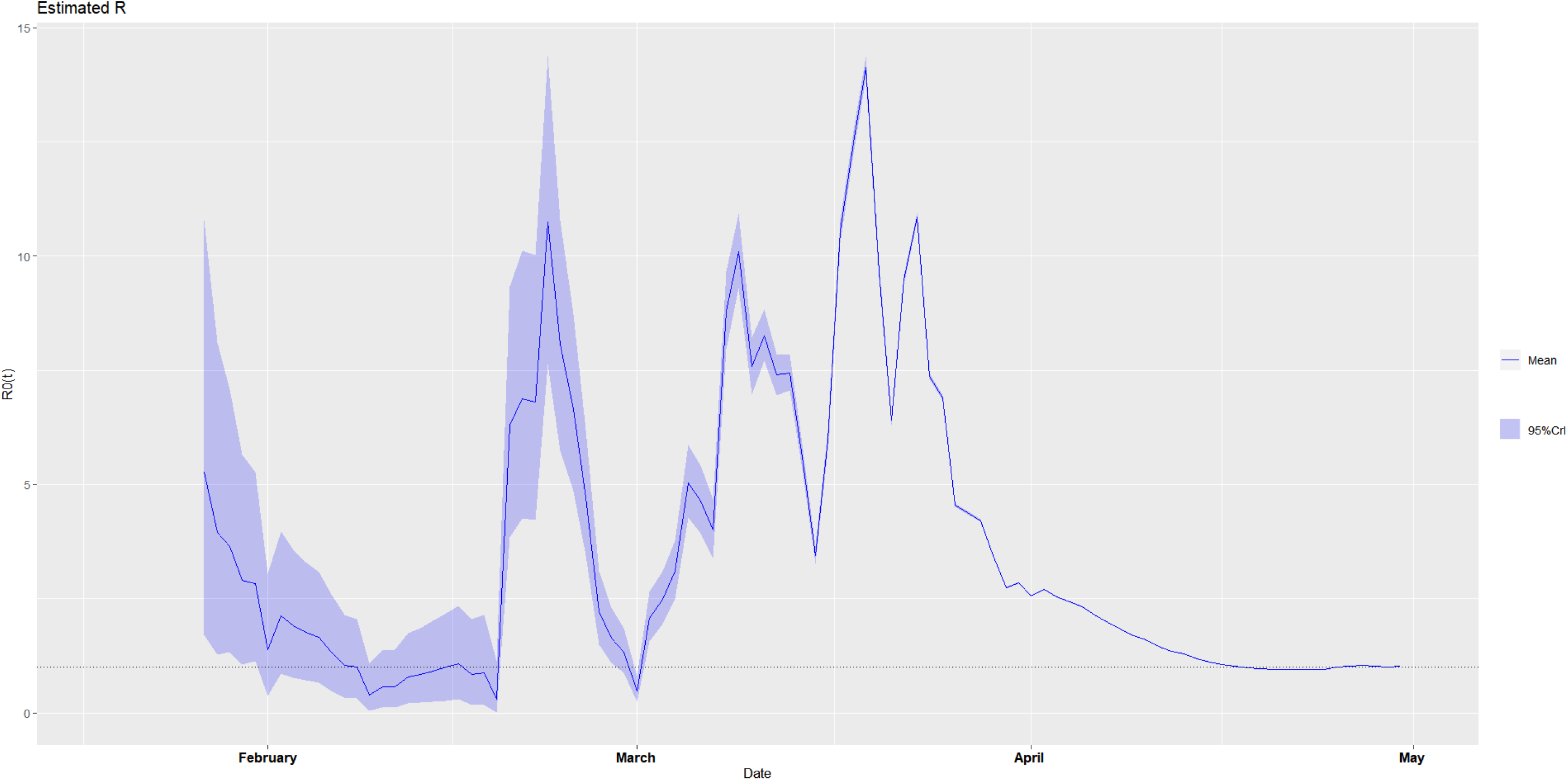
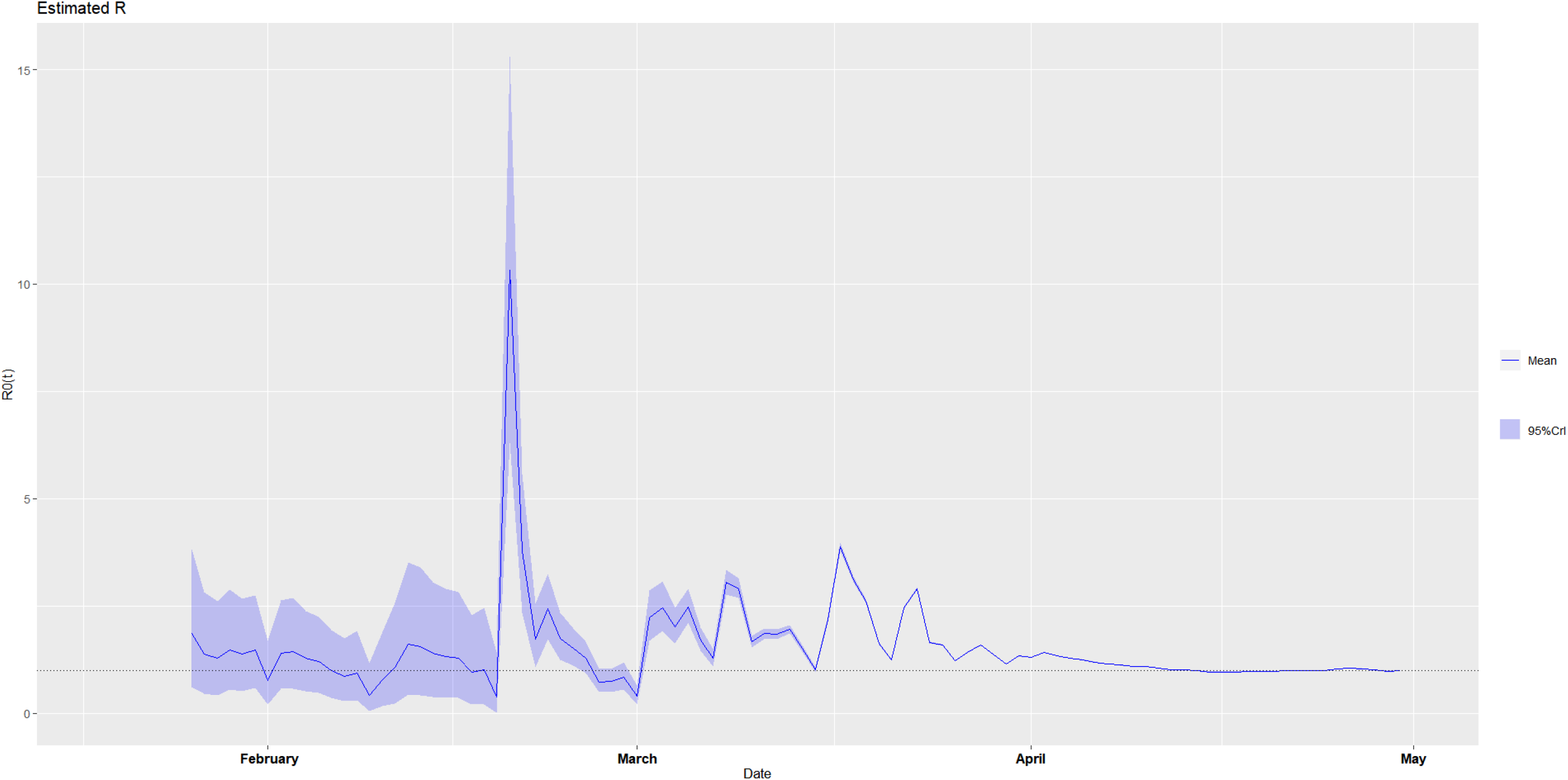
The estimated time-varying reproduction number during the ongoing pandemic of the coronavirus disease 2019 (COVID-19) in the United States of America from January 19, 2020 to April 30, 2020 under two scenarios. **A. Scenario 1:** We specified the mean (SD) of the Gamma distribution of serial interval (SI) to be 8.4 (3.8) days to mimic the 2003 epidemic of the severe acute respiratory syndrome (SARS) in Hong Kong.^7,8^ **B. Scenario 2:** We specified the mean (SD) of the Gamma distribution of serial interval (SI) to be 2.6 (1.5) days to mimic the 1918 pandemic of influenza in Baltimore, Maryland.^7,8^

**Figure 12-1.**
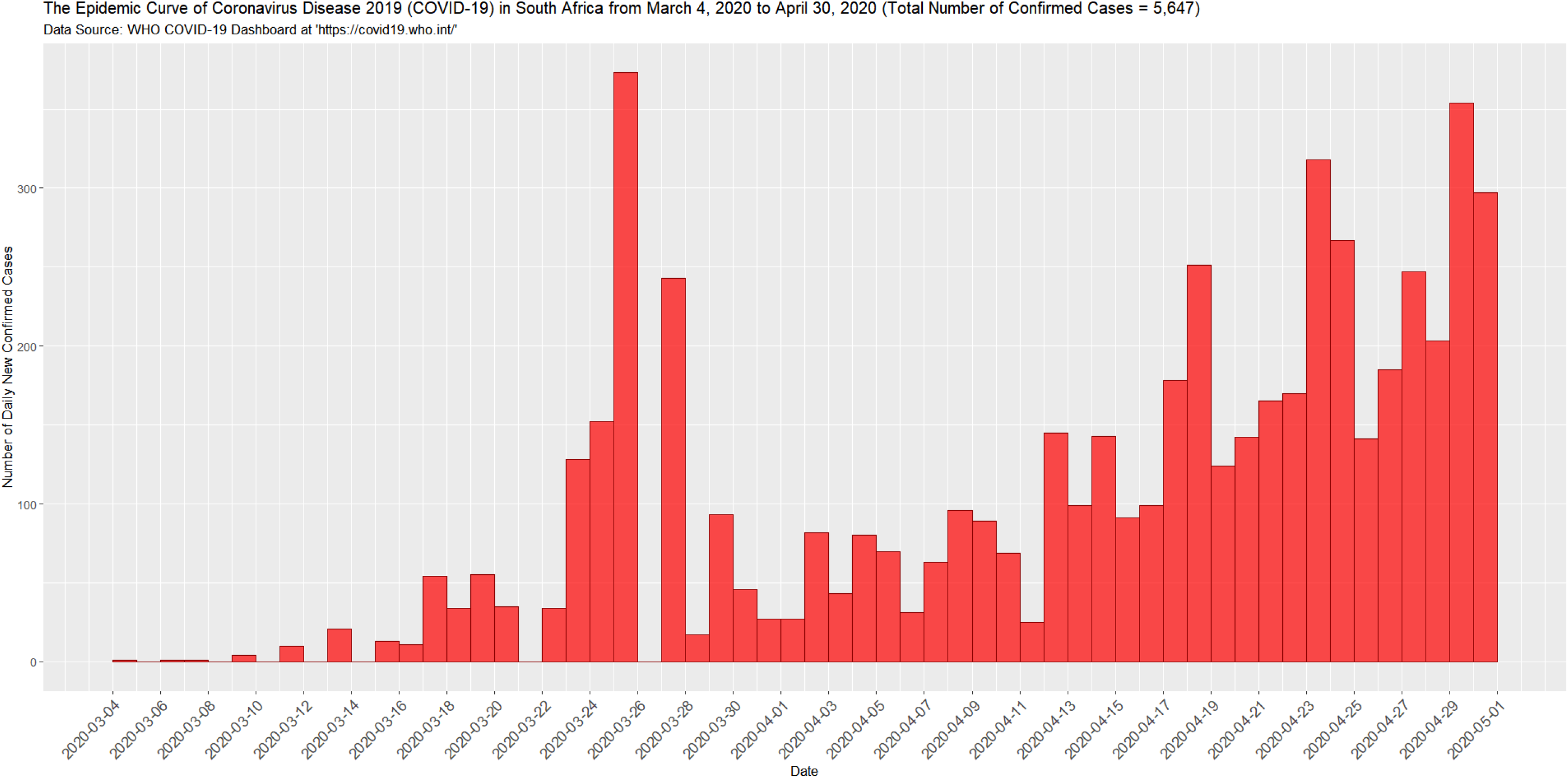
The epidemic curve of the coronavirus disease 2019 (COVID-19) in South Africa from March 4, 2020 to April 30, 2020.

**Figure 12-2.**
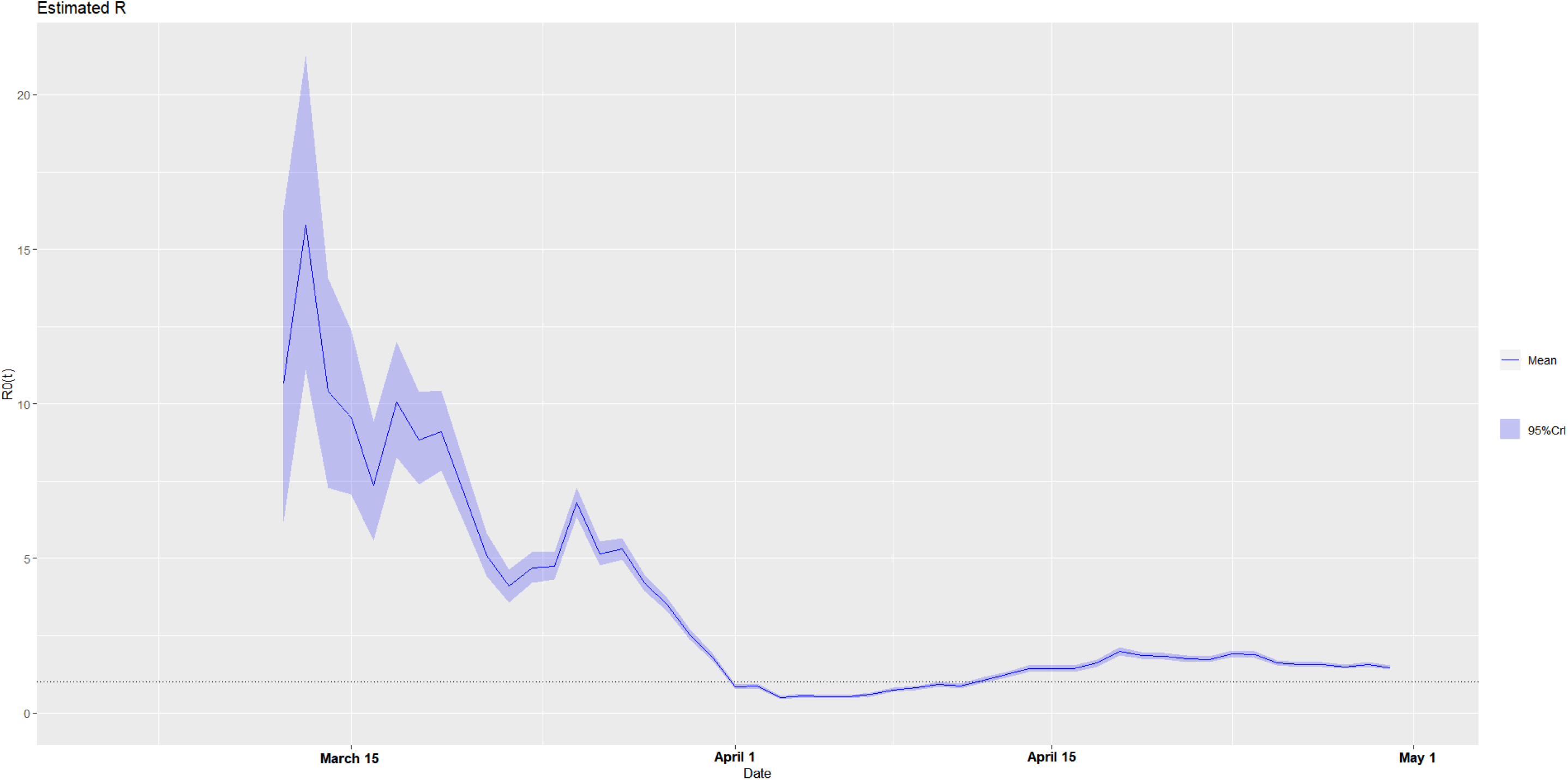
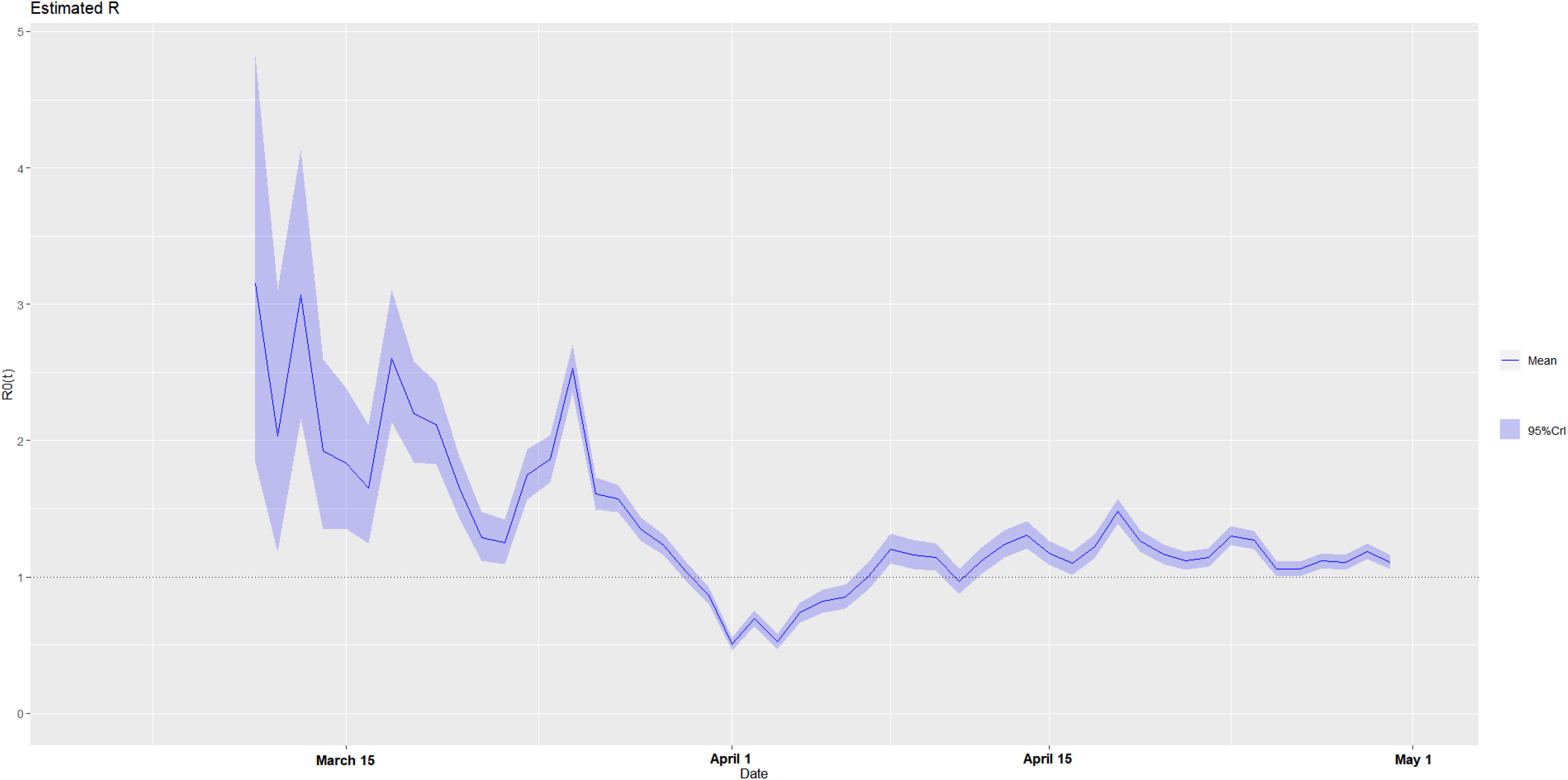
The estimated time-varying reproduction number during the ongoing pandemic of the coronavirus disease 2019 (COVID-19) in South Africa from March 4, 2020 to April 30, 2020 under two scenarios. **A. Scenario 1:** We specified the mean (SD) of the Gamma distribution of serial interval (SI) to be 8.4 (3.8) days to mimic the 2003 epidemic of the severe acute respiratory syndrome (SARS) in Hong Kong.^7,8^ **B. Scenario 2:** We specified the mean (SD) of the Gamma distribution of serial interval (SI) to be 2.6 (1.5) days to mimic the 1918 pandemic of influenza in Baltimore, Maryland.^7,8^

Finally, we listed each country’s estimated distributional parameters of *R*_0_(*t*) for the weekly window ending on April 30, 2020 under the two plausible scenarios (**A** and **B**) in Table 2 as a numerical complement to Figures 1-2A and 1-2B, 2-2A and 2-2B, …, and 12-2A and 12-2B. The values of the posterior mean and median were close to each other during the study period. The bigger the size of the SD, the more the number of daily new confirmed cases in the last week of April. Intriguingly, after comparing the posterior mean values between the two plausible scenarios (**A** and **B**), we found that (1) A > B: Singapore, the United States of America, and South Africa, and (2) B > A: all the other 9 countries. It seemed reasonable to take an average of these two as an approximate *R*_0_(*t*) for comparison. By doing so, the United States of America (**A**: 1.0197, **B**: 0.9974) and South Africa (**A**: 1.4655, **B**: 1.1080) were the two countries with the approximate *R*_0_(*t*) ≥ 1.0 at the end of April, and thus they were now facing the harshest battles against the coronavirus among the 12 selected countries. By contrast, Spain (**A**: 0.4574, **B**: 0.6892), Germany (**A**: 0.5055, **B**: 0.7023), and France (**A**: 0.5092, **B**: 0.7340) with smaller values of the estimated *R*_0_(*t*) were relatively better than the other 9 countries.

**Table 2.** The estimated distributional parameters of the time-varying reproduction number for the weekly window ending on April 30, 2020 during the ongoing pandemic of the coronavirus disease 2019 (COVID-19) in the 12 selected countries outside China from January 11, 2020 to May 1, 2020 under two scenarios. **A. Scenario 1:** We specified the mean (SD) of the Gamma distribution of serial interval (SI) to be 8.4 (3.8) days to mimic the 2003 epidemic of the severe acute respiratory syndrome (SARS) in Hong Kong.^7,8^ **B. Scenario 2:** We specified the mean (SD) of the Gamma distribution of serial interval (SI) to be 2.6 (1.5) days to mimic the 1918 pandemic of influenza in Baltimore, Maryland.^7,8^

| Country | Scenario | Mean | SD | Quantile |  |  |  |  |  |  |
| --- | --- | --- | --- | --- | --- | --- | --- | --- | --- | --- |
|  |  |  |  | 0.025 | 0.05 | 0.25 | 0.50 | 0.75 | 0.95 | 0.975 |
| 1. Singapore | A | 0.8962 | 0.0127 | 0.8715 | 0.8754 | 0.9171 | 0.8961 | 0.9171 | 0.9171 | 0.9212 |
|  | B | 0.8209 | 0.0116 | 0.7983 | 0.8019 | 0.8401 | 0.8209 | 0.8401 | 0.8401 | 0.8438 |
| 2. South Korea | A | 0.5414 | 0.0677 | 0.4169 | 0.4351 | 0.6573 | 0.5386 | 0.6573 | 0.6573 | 0.6819 |
|  | B | 1.0017 | 0.1224 | 0.7763 | 0.8093 | 1.2112 | 0.9968 | 1.2112 | 1.2112 | 1.2554 |
| 3. Japan | A | 0.5891 | 0.0135 | 0.5629 | 0.5670 | 0.6116 | 0.5890 | 0.6116 | 0.6116 | 0.6160 |
|  | B | 0.7794 | 0.0179 | 0.7447 | 0.7502 | 0.8091 | 0.7793 | 0.8091 | 0.8091 | 0.8149 |
| 4. Iran | A | 0.6883 | 0.0086 | 0.6716 | 0.6742 | 0.7024 | 0.6882 | 0.7024 | 0.7024 | 0.7052 |
|  | B | 0.8267 | 0.0103 | 0.8067 | 0.8099 | 0.8437 | 0.8267 | 0.8437 | 0.8437 | 0.8470 |
| 5. Italy | A | 0.6303 | 0.0054 | 0.6198 | 0.6215 | 0.6392 | 0.6303 | 0.6392 | 0.6392 | 0.6409 |
|  | B | 0.7658 | 0.0066 | 0.7530 | 0.7551 | 0.7766 | 0.7658 | 0.7766 | 0.7766 | 0.7787 |
| 6. Spain | A | 0.4574 | 0.0046 | 0.4484 | 0.4499 | 0.4649 | 0.4574 | 0.4649 | 0.4649 | 0.4664 |
|  | B | 0.6892 | 0.0069 | 0.6758 | 0.6779 | 0.7006 | 0.6892 | 0.7006 | 0.7006 | 0.7028 |
| 7. Germany | A | 0.5055 | 0.0054 | 0.4949 | 0.4966 | 0.5144 | 0.5054 | 0.5144 | 0.5144 | 0.5161 |
|  | B | 0.7023 | 0.0075 | 0.6876 | 0.6900 | 0.7147 | 0.7022 | 0.7147 | 0.7147 | 0.7171 |
| 8. France | A | 0.5092 | 0.0059 | 0.4977 | 0.4995 | 0.5189 | 0.5091 | 0.5189 | 0.5189 | 0.5207 |
|  | B | 0.7340 | 0.0085 | 0.7175 | 0.7201 | 0.7480 | 0.7340 | 0.7480 | 0.7480 | 0.7507 |
|  |  |  |  | 0.025 | 0.05 | 0.25 | 0.50 | 0.75 | 0.95 | 0.975 |
| 9. Belgium | A | 0.5901 | 0.0083 | 0.5739 | 0.5765 | 0.6038 | 0.5900 | 0.6038 | 0.6038 | 0.6064 |
|  | B | 0.7585 | 0.0107 | 0.7377 | 0.7410 | 0.7761 | 0.7584 | 0.7761 | 0.7761 | 0.7795 |
| 10. United Kingdom | A | 0.7746 | 0.0047 | 0.7654 | 0.7669 | 0.7823 | 0.7746 | 0.7823 | 0.7823 | 0.7838 |
|  | B | 0.8376 | 0.0051 | 0.8277 | 0.8292 | 0.8460 | 0.8376 | 0.8460 | 0.8460 | 0.8476 |
| 11. United States of America | A | 1.0197 | 0.0023 | 1.0153 | 1.0160 | 1.0234 | 1.0197 | 1.0234 | 1.0234 | 1.0241 |
|  | B | 0.9974 | 0.0022 | 0.9931 | 0.9938 | 1.0011 | 0.9974 | 1.0011 | 1.0011 | 1.0018 |
| 12. South Africa | A | 1.4655 | 0.0356 | 1.3966 | 1.4075 | 1.5246 | 1.4653 | 1.5246 | 1.5246 | 1.5361 |
|  | B | 1.1080 | 0.0269 | 1.0559 | 1.0641 | 1.1526 | 1.1078 | 1.1526 | 1.1526 | 1.1614 |

Abbreviation: SD = standard deviation.

## Discussion

Almost everyone is susceptible to the novel COVID-19 and this is one of the reasons why the COVID-19 epidemic occurs in many places and has caused public panics worldwide. In terms of population size, the depletion due to death or recovery might be negligible in big countries such as China, but not in small ones such as Singapore. Many recoveries are probably not susceptible any more in the late phase of the epidemic. Although there was officially no imported case of COVID-19 in China at the beginning of the COVID-19 epidemic, the imported cases of COVID-19 are the major sources of initial infection in many other countries starting from February of 2020. These features made the task of modeling this epidemic in any country outside China more difficult. Moreover, since the COVID-19 is new to human society, its diagnostic criteria, control measures, and medical care are inevitably changing during the pandemic as the knowledge and experience about it are accumulated continuously.^4,5^ Thus, the result obtained in this study is merely a rough estimate of *R*_0_(*t*) for each of the 12 selected countries. Nevertheless, our findings for the 12 selected countries passing the early phase of the pandemic are consistent with the early estimates of *R*_0_ for the COVID-19 epidemic such as 2.24 (95% confidence interval [CI]: 1.96–2.55)–3.58 (95% CI: 2.89–4.39),^9^ 2.2 (95% CI: 1.4–3.9),^10^ 2.6 (uncertainty range: 1.5–3.5),^11^ 2.68 (95% CrI: 2.47–2.86),^12^ 1.4–2.5 (WHO),^13^ 2.0–3.3,^13^ 2.2 (90% CI: 1.4–3.8),^13^ 6.47 (95% CI: 5.71–7.23),^13^ 3.11 (90% CI: 2.39–4.13),^13^ and 2.0.^13^

This study had several limitations because we relied on some assumptions to make a rapid analysis of this ongoing epidemic feasible. First, we assumed that all new cases of COVID-19 in each of the 12 selected countries are detected and reported to the WHO correctly. However, asymptomatic or mild cases of COVID-19 are likely undetected, and thus under-reported, especially in the early phase of the epidemic.^5,14^ In some countries, a lack of diagnostic test kits for the SARS-CoV-2 and a shortage of qualified manpower for fast testing can also cause under-reporting or delay in reporting. Nevertheless, we only analyze the official source of epidemic data for examining the trend of *R*_0_(*t*) during an epidemic because it is feasible, transparent, and reproducible. Unintentional under-reporting is inevitable in any country during an ongoing epidemic due to various reasons. Second, since the reported number of daily new confirmed cases includes both domestic and repatriated cases according to the description of WHO,^4^ the imported cases lead to an over-estimation of *R*_0_(*t*) in our epidemic analysis. Yet, they most likely appear in the early phase of the COVID-19 pandemic, and then have less effect on the estimated *R*_0_(*t*) in the later phase of the pandemic. The estimate_R function of the EpiEstim package provides the option for including the separated data of imported cases in the estimation of *R*_0_(*t*).^7^ Thus, our epidemic analysis can be easily refined once the domestic and repatriated cases are reported separately in the future. Third, we admitted the time delay in our estimates of *R*_0_(*t*) for the COVID-19 pandemic in the 12 selected countries due to the following two time lags: (1) the duration between the time of infection and the time of symptom onset (i.e., the *incubation period* of infection) if infectiousness began around the time of symptom onset^8^ and (2) the duration between the time of symptom onset and the time of diagnosis.^5^ Nevertheless, since asymptomatic carriers can transmit the COVID-19,^15-17^ the first time lag is actually shorter than expected and it becomes the duration between the time of infection and the time of becoming infectious (i.e., the *latent period* of infection). Moreover, the time interval from symptom onset to diagnosis is shorter and shorter due to the full alert of society and the faster diagnostic tests.^5^ Fourth, we assumed that the distribution of SI does not change considerably over time as the pandemic progresses. However, as for the SARS epidemic in Singapore, the SI tended to be shorter after control measures were implemented.^18^ The SI also depended on the amount of infecting dose, the level of host immunity, and the intensity of person-to-person contacts.^16^ To summarize, most of these limitations led our estimation of *R*_0_(*t*) into a more conservative context.

Looking at the epidemic curves of the 12 selected countries in Figures 1-1, 2-1,…, 12-1 respectively, we could see that the size of the epidemic varies across these 12 countries. It primarily depends on how early and how fast the effective control measures are taken to stop the spread of the coronavirus from the very beginning of the COVID-19 pandemic. Any mistakes in this matter would have a dramatic impact on the sequel. The commonly used lag-1 relative change (%) in the number of daily new confirmed cases is not useful because its denominator varies and the time lag is not long enough to reflect what is really going on behind those numbers.

Next, as shown in Figures 1-2A and 1-2B, 2-2A and 2-2B,…, and 12-2A and 12-2B respectively, the estimated *R*_0_(*t*) over sliding weekly windows for each of the 12 selected countries begins to drop gradually after a certain time period. Yet, when the number of new confirmed cases is climbing up day by day before it reaches the peak(s), everyone is keen to know: When will the epidemic curve reach the hilltop?^19^ Although it is difficult to estimate the exact date when it will happen, the peak can only appear after the trend of the computed *R*_0_(*t*) is declining sharply and monotonically for some days (e.g., about 10 days in China), and then the daily number of new confirmed cases will begin dropping, indicating that the COVID-19 epidemic has abated. However, it is very frustrating to see the curve of the estimated *R*_0_(*t*) staggering slowly down to the threshold line of *R*_0_(*t*) = 1.0 in most of the 12 selected countries. Notice that the same value of *R*_0_(*t*) may have quite different magnitudes of impacts on the number of daily new confirmed cases, depending on how many infectious cases in the community at the time. Even though *R*_0_(*t*) = 1.0, the number of daily new confirmed cases can still go up soon if more and more infectious subjects are accumulated in society. Thus, a steeply sloped line of the estimated *R*_0_(*t*) is crucial — that is, how early and how fast does it go downhill? Finally, the COVID-19 epidemic in China ended around March 7-8, 2020, indicating that the prompt and aggressive control measures of China were very effective,^20-23^ but when can it happen in the 12 selected countries? In particular, the resurgence has occurred in Singapore, South Korea, Japan, Iran, and South Africa respectively. Thus, when the COVID-19 pandemic will end in any of the 12 selected countries and the whole world worries us. Given a symmetric one-peak epidemic curve, the total time and size of the epidemic is just the double of the time and size from the beginning of the epidemic to its peak. As compared to China’s experience of success, each of the 12 selected countries should quickly learn from the fresh lessons of the past two to three months, and then find much more effective ways to fighting the COVID-19 pandemic for winning the battle against the coronavirus (see Appendix). In our opinion, seeing the estimated *R*_0_(*t*) going downhill speedily is more informative than looking for the drops in the daily number of new confirmed cases during an ongoing epidemic of infectious disease. And, the consistent decline of the estimated *R*_0_(*t*) in trend over time is more important than the actual values of the estimated *R*_0_(*t*) themselves. The steeper the slope, the sooner the epidemic ends.

Finally, we urged public health authorities and scientists worldwide to estimate time-varying reproduction numbers routinely during epidemics of infectious diseases and to report them daily on their websites such as the “Tracking the Epidemic” of China CDC for guiding the control strategies and reducing the unnecessary panic of the public until the end of the epidemic.^23^ Fitting complex transmission models of infectious disease dynamics to epidemic data with limited information about required parameters is a challenge.^6,8,24-26^ The results of such analyses may be difficult to generalize due to the context-specific assumptions made and it can be too slow to meet a pressing need during an epidemic.^8,18,27-34^ Thus, an easy-to-use pragmatic tool such as the estimated *R*_0_(*t*) for monitoring the COVID-19 pandemic is so important in practice. Although this study could not provide the most accurate results rigorously, it sufficed for the pragmatic purpose from the public health viewpoint. We believed that it was an approximate answer to the right question timely. Refinements in the estimation of *R*_0_(*t*) can be made with the individual patient data, including personal contact history, whenever they are available for analysis.

Since the coronavirus has spread out globally, we should take the important lessons from China,^2,20-23,31-34^ South Korea,^35^ Italy,^36^ the United States of America,^37,38^ and Singapore^39^ and learn the experiences from the previous epidemics^1,25^ to mitigate its harm as much as possible. We may also use China as an example to anticipate the potential progression of the COVID-19 pandemic in a particular country.^2^ As listed in Appendix, control tactics and measures should be applied in line with local circumstances,^25^ but the same easy-to-use monitoring tool, *R*_0_(*t*), can be applied to many places. Such timely available information for monitoring the ongoing VOVID-19 pandemic deserves societal attention, especially when the government is going to make a move in the harsh battle against the coronavirus. As the coronavirus outbreak continues to spread, let’s help each other to combat the COVID-19 pandemic together. After all, we are all in the same shaking boat now.

## Data Availability

The publicly available COVID-19 pandemic data can be downloaded from the "WHO COVID-19 Dashboard" website (https://covid19.who.int/).

## Appendix: The source-transmission-host approach to controlling the COVID-19 pandemic

### 1. Goal: The “Separation” Process

As depicted in the following diagram, what we should do to control the COVID-19 epidemic within a black rectangle (e.g., a city, a state, or a country) is to *separate* the red balls (i.e., patients) from the grey circles (i.e., medical personnel) and the green balls (healthy subjects). When the number of new cases is soaring in Stage 2, the more aggressive *separation process* is needed by reducing population mobility, implementing mass testings, and conducting community surveillance. We ask or force the red balls to enter the big grey circle(s) as in Stage 3.

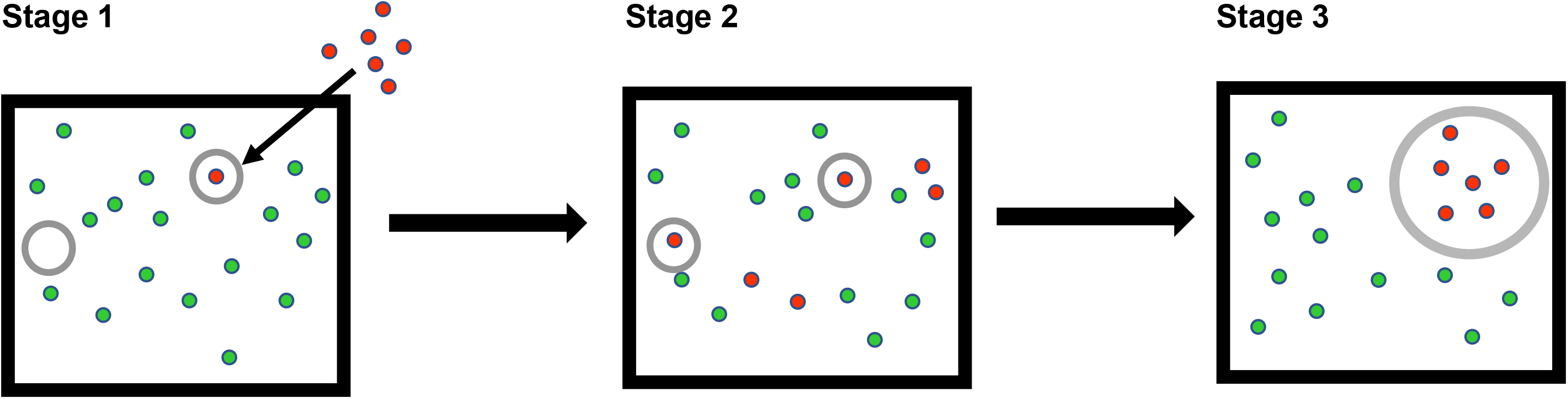

### 2. Control Tactics and Measures: The “Source → Transmission → Host” Approach

2. Control Tactics and Measures: The “Source → Transmission → Host” Approach.
| Priority<sup>1</sup> | Methods |  |  |
| --- | --- | --- | --- |
|  | Source | Transmission | Host |
| 1 | Diagnosis<sup>2</sup> (e.g., traveling history,<sup>3</sup> contact history,<sup>3</sup> symptoms, fast testing, and X-ray examination) | Local, national, or international traveling restrictions | Health education through news and social media (esp., the strategies for fighting the COVID-19 pandemic and risk communication) |
| 2 | Isolation of laboratory-confirmed cases | Contact tracing for quarantines | Hand washing |
| 3 | Isolation of suspected cases<sup>4</sup> | Quarantines for at least 14 days | Face-mask wearing<sup>1</sup> |
| 4 | Medical treatments<sup>5</sup> (Not available yet, but recent studies show that proper uses of chloroquine / hydroxychloroquine and remdesivir may be effective) | Epidemic monitoring (e.g., estimating the fixed-time and time-varying basic reproduction number, $R_0$ and $R_0(t)$) | Contact reductions (e.g., stay at home, working from home, video conferences, distance learning, telemedicine, and shopping online) |
| 5 | Supportive cares | Social distancing<sup>6</sup> | Balanced nutrition |
| 6 | Disinfection and decontamination | School closing | Regular exercises |
| 7 | Contact tracing for identifying the source of infection | Social disruption (e.g., limitations in social gatherings and entertainment activities, stay-at-home orders, and business shutdowns) | Good sleep |
| 8 | Community-based surveillances | Traveling bans | Peaceful and proactive mind |
| 9 | Mass testings or aggressive | Border closing | Indirect social contacts to care and |

|  | screening |  | help each other |
| --- | --- | --- | --- |
| 10 | Preparedness of medical facilities, personnel, protective equipment, medical supplies, and contingency plan | Curfews (short-term or long-term) | Medical treatments<sup>5</sup> (Not available yet, but recent studies show that proper uses of chloroquine / hydroxychloroquine and remdesivir may be effective), isolation, and supportive cares for laboratory-confirmed cases |
| 11 | Reporting and announcements (e.g., risk sources, locations, and pathways) | Lockdowns (short-term or long-term) | Vaccination<sup>5</sup> (Not available yet, but scientists are developing COVID-19 vaccines now) |
| 12 |  | Information release and sharing (e.g., risk sources, locations, and pathways) | Long-term follow-ups of recovered cases |

- <sup>1</sup> Control tactics and measures should be applied in line with local circumstances at various stages (i.e., Stages 1, 2, and 3) in the global battle against the coronavirus (SARS-CoV-2). For example, wearing medical face masks is recommended to everyone only in Stage 2, where no one in a community is very sure whether he/she may get an infection or he/she is infectious or not. - <sup>2</sup> The diagnostic criteria have been changing over time as the experience and knowledge are accumulated continuously during the ongoing COVID-19 pandemic. - <sup>3</sup> In checking traveling history, indirect contacts with someone from the infected country at the place outside both countries should also be noticed. - <sup>4</sup> Suspected cases, including clinically diagnosed cases, are the subjects who are waiting for the testing results. - <sup>5</sup> Medical treatments and vaccination are the *pharmaceutical interventions* (PI), whereas the other control tactics and measures are the *nonpharmaceutical interventions* (NPI). - <sup>6</sup> Social distancing should consider not only the observable physical distance between two subjects but also the seemingly unobservable distance in the social networks of the two subjects.

